# Development of a Core Outcome Measure Instrument; “LeishCOM_LCL”, for Localised Cutaneous Leishmaniasis

**DOI:** 10.1101/2024.05.28.24307884

**Authors:** Shalindra Ranasinghe, Sujai Senarathne, Vijani Somaratne, Charles JN Lacey, Surangi Jayakody, Amila Wickramasinghe, Indira Kahawita, Hiro Goto, Mitali Chatterjee, José AL Lindoso, Vivak Parkash, Surya J Chaudhuri, Renu Wickremasinghe, Nilay K. Das, Paul M. Kaye, Alison M Layton

## Abstract

**Background:** Localized cutaneous leishmaniasis (LCL) is a chronic ulcerating disease. A literature review identified inconsistencies in clinical trials. The aims of this study were to reach a consensus on the most important domains to measure when assessing LCL, agree on parameters to measure the domains, and develop a tool representing a Core Outcome Set (COS), for use in clinical assessment of LCL.

**Methodology & Principal findings:** A literature review was conducted to identify any existing COS for LCL embracing agreed Outcome Domains, i.e. what to measure and any Outcome Measurement Instruments (OMIs). As no COS was available, potential outcome domains for assessment of LCL were identified through an international collaborative approach using e-consultations and virtual discussions with expert stakeholders (n=20) from geographically different LCL endemic countries. Subsequent judgmental validation process included a face-to-face multidisciplinary stakeholders’ meeting adopting the Nominal Group Technique. A final consensual agreement on outcome domains and items required to measure these domains was established. “Clinical Cure” was defined as the ideal overall "General Concept". The five Core Outcome Domains included **Signs** capturing clinical morphology, diameter, and induration of an index lesion with the aid of a palpability score, **Treatment Efficac**y assessing percentage change in size of the lesion and re-epithelialization compared to baseline, **Treatment Impact** which included an investigator and patient visual analogue score, and **Clinical Sequelae** rating pigment change, atrophic and hypertrophic/keloid scars. It was agreed that two open-ended questions should be included to capture some aspects of **Health-Related Quality of Life** as a means of capturing a patient-focused approach.

**Conclusion:** LeishCOM_LCL was generated to reflect a COS for LCL. This captured demographic details, agreed outcome domains and measures to assess these domains. Validation of LeishCOM_LCL will be reported in a separate paper. Development of a Patient Reported Outcome Measure will be considered in the future.

**Author Summary:** Localized cutaneous leishmaniasis (LCL) is a chronic ulcerating disease caused by the parasite *Leishmania* spp. Literature review identified inconsistencies in methods and parameters used to evaluate treatment/alternative-interventions resulting in difficulties in comparing new treatment/interventions in clinical trials. In our international consensual study, we adopted the face-to-face nominal group technique and a judgment process to identify domains key to assessment of LCL. Subsequent measures for each domain were used to form a Core Outcome Set (COS). LeishCOM_LCL was developed as an outcome measure instrument (OMI) to capture the COS incorporating existing and newly developed tools. “Clinical Cure” was agreed as the “General Concept” to be captured through five domains. It was agreed that “Signs” domain should capture clinical morphology, diameter, and induration of an index lesion with the aid of a palpability score. “Treatment Efficacy” was assessed by recording percentage change in size of the lesion and re-epithelialization compared to baseline. “Treatment Impact” was reflected through an investigator and the patient visual analogue score and “Clinical Sequelae” rated pigment change, atrophic/hypertrophic scars. Two open-ended questions were included to capture some aspects of “Health-Related Quality of Life”. CutLeishCOM also records patient demographic details and was validated in a small cohort of patients.

## Introduction

Localized cutaneous leishmaniasis (LCL) is a skin disease caused by an intra-cellular protozoan parasite belonging to the genus *Leishmania* that is transmitted through a bite of an infected female phlebotomine sand fly. It is considered a neglected tropical disease and is endemic in 90 countries with an estimated 1 million new cases reported annually [1]. LCL is usually characterized by the presence of amastigotes localized in skin tissue. It appears in exposed areas of the body and frequently heals with lifelong scars [1]. This form of presentation does not include mucosal lesions and associated disseminated / diffuse CL or Post Kala-azar Dermal Leishmaniasis [1, 2]. The skin lesions are typically chronic in nature and the disease shows a wide range of clinical features ranging from a small papule to extensive ulceration and can often result in permanent physical and psychological sequelae with a potentially life-long impact. Clinicians frequently treat LCL with the aim of minimizing sequelae such as scarring that may result in disfigurement and social stigma [3]. Treatment modalities available for LCL include intra-lesional or parenteral pentavalent antimonial compounds and liposomal amphotericin B as the mainstay of treatment in Old World LCL while miltefosine and pentamidine are also used in the treatment of New World LCL [4-7]. Cryotherapy and thermotherapy are some of the commonly used non-pharmacological treatment measures [7].

A review of the clinical trials examining different treatment modalities for LCL found studies to be deficient in design, execution, analysis, and reporting [8]. Furthermore, systematic literature reviews demonstrate that most clinical trials on LCL fail to clearly identify consistent and standardized primary and secondary clinical outcome measures [9-12]. There are a few new treatment/alternative-interventions described for LCL [7,13-15]. However, the results of those studies are difficult to compare due to inconsistencies in the methods and the parameters used for the evaluation. Furthermore, most studies have not considered the patients’ perspective. Recommendations to assess initial response and define timelines to initial clearance and subsequent cure have been suggested as potential important outcomes [16, 17]. The development of a validated scoring system for LCL based on harmonized methodologies would allow assessment of treatment response in routine clinical settings and enable comparison of the efficacy of existing or new drugs as well as novel alternative interventions and would subsequently support meta-analysis.

Core Outcome Sets (COS) represent agreed standardized outcomes that should be reported for all trials conducted in a specific research area, with the intention of reducing bias and ensuring that data from different trials are suitable for meta-analysis [18]. Various COS which embrace agreed Core Outcome Domains and Core Outcome Measures to assess the domains have been developed for several dermatological disorders as a means of evaluating the severity and impact of the condition as well as therapeutic response to treatment in a standardized manner. The development of clinical scores through measurable outcomes using clinimetrics and inclusion and scoping out of clinicians’ and patients’ perspectives results in improved outcome measures [19, 20].

Many measures have been developed based on the cardinal clinical features of each disorder, e.g. the Psoriasis Area and Severity Index (PASI) [21], the Eczema Area and Severity Index (EASI) [22] and the Vitiligo Area Scoring Index (VASI) [23] etc. These instruments only account for the clinical severity of the disease and treatment response but fail to capture the clinical sequelae and the impact of the disease on the quality of life. The Harmonizing Outcome Measures for Eczema (HOME) roadmap was developed and implemented in cooperation with the COMET (Core Outcome Measures in Effectiveness Trials) and COSMIN (COnsensus-based Standards for the selection of health Measurement INstruments) research groups, with the aim of developing a standardized, validated and consensus-based roadmap for developing COS for atopic eczema [24]. The process developed for the HOME roadmap has been recommended and adopted for other dermatological conditions [19, 24]. However, this approach has rarely been adopted for neglected diseases of the skin.

There are currently only two studies that report a harmonized approach to assess LCL, one describing harmonized measurable clinical methodologies to assess the response to interventions in clinical trials [16] and a second follow-up study [25] assessing the capacity of implementation of the harmonized methodologies across several geographic regions. However, the proposed measurable outcomes in these studies have not been validated. In addition, Patient Reported Outcome Measures (PROMs) that qualitatively assess the impact of the disease and / or the response to treatment or adverse effects from therapy were not considered. The absence of sequelae resulting from LCL is a further gap in current assessments [16, 25]. Therefore, the aim of our study was to reach a consensus on the most important Core Outcome Domains that need to be considered and measured when assessing LCL and to further develop and validate clinical measures as a part of an overall assessment tool to capture the response to treatment in LCL.

The overall aim of this work was to adopt a standardized and validated approach to assess agreed clinical aspects of LCL and response to treatment in a measurable manner for use in clinical trials and routine patient care which were then reflected in a practical tool. This paper describes identification through consensus of what to measure in LCL clinical trials (Core Outcome Domains) and how to measure these aspects as well as the process involved in the development of a clinical instrument which captures and measures the areas identified. Once developed, the Leishmaniasis Core Outcome Measure Instrument for Localised Cutaneous Leishmaniasis (LeishCOM_LCL) was incorporated into a case report form (CRF) and the process of face and content validation was undertaken. Further comprehensive validation of the final outcome measure instrument has been conducted through a clinical study. The results from this further validation along with the methodology and detailed data will be reported in a separate manuscript (manuscript under preparation).

## Methods

### Ethical clearance

Ethical approval for the study was obtained from the University of Sri Jayewardenepura, Sri Lanka to develop a Core Outcome Measure Instrument aligned to HOME methodology and this included ethical approval to enroll patients for the face validity and future validation of any instrument developed (ERC 52/17).

### Identification of Core Outcome Measures

Core Outcome Domains and Measures to assess agreed Domains were identified to inform an Outcome Measure Instrument (OMI; LeishCOM_LCL) for LCL. The HOME methodological framework was adopted using the following steps:

#### 1. Development process; define scope and applicability

A comprehensive literature review was carried out to identify publications that had identified COS including domains, measures and instruments already aligned to LCL using PubMed, MEDLINE, Cochrane library, COMET initiative, and COSMIN websites. This review identified the already published work on harmonized outcome measures in LCL [16, 17, 25]. The principal investigators laid down the conceptual framework. Formalizing the Core Outcome Domains for LCL had not been established and this was therefore taken forward through a collaborative international approach involving virtual discussions and e-consultations with stakeholder clinicians including dermatologists and their teams who care for LCL from Sri Lanka, India, Brazil and the United Kingdom.

#### 2. Judgment process; define core set of outcome domains

A subsequent judgment process was then undertaken adopting the nominal group technique (NGT) to reach a final consensual agreement on the set of Core Outcome Domains. The NGT approach was performed by having face-to-face discussions at a workshop in March 2018 in Sri Lanka. The NGT approach was selected to ensure that there was an opportunity to share clinical experience and secure clear consensus among stakeholders. A multi-institutional, multi-disciplinary stakeholder panel of international experts from Sri Lanka, India, Brazil, and the United Kingdom comprising of dermatologists, general physicians and parasitologists were included along with a moderator (n= 20). For each Core Outcome Domain, a review of previous approaches to measure the domain was considered [16, 17, 25]. During the workshop, items to measure the domains were also evaluated and multiple rounds of discussions were carried out to secure agreement within the panel of experts on "what to measure" which informed the final Core Outcome Domains as well as “how to measure” the domains. Patient perspectives including clinical and psychological aspects were captured by doing a field visit by the expert stakeholders one day before the NGT meeting to a hospital-based dermatology clinic in a LCL endemic area (Hambantota) in Sri Lanka and had face-to-face discussions with LCL patients. Also, the international clinicians taking care of their LCL patients in other countries presented additional clinical and psychological perspectives within their region to ensure that the most important patients’ perspectives encountered in different endemic areas were captured.

As noted previously, for each Core Outcome Domain, previous approaches on "how to measure the domains" were considered. The proposed items to measure the domains were then evaluated by multiple rounds until the same panel of experts reached a consensus. Once established, the construction of a novel clinical assessment tool "LeishCOM_LCL" for practical use in the field was developed. The various measures that informed the tool "LeishCOM_LCL" underwent multiple reviews during the period of reaching consensus and this included practical approaches to ensure that the assessments were conducted in a standardized manner.

Furthermore, during this process, a subjective judgment of the content validity was done by assessing: i) the degree to which no important items were missing (comprehensiveness), ii) the degree to which the items were correctly understood by the clinician and the patient (comprehensibility), and iii) relevance of the content of OMI for the assessment of healing of LCL lesions. The final version of the tool was incorporated into a case report form (CRF) for downstream application when assessing LCL patients in the field and during this process underwent some face validity with more robust validation which will be described and reported in a separate manuscript.

#### 3. Case Report Form (CRF) for data collection

The case report form (CRF) allowed data collection and validation of LeishCOM_LCL. It captures demographic details, reflects the core outcome domains, agreed outcome measurements, and includes scoring systems for the selected COS including visual analogue scores that capture clinician’s and patient’s perspectives. The CRF also includes two open-ended questions aimed at capturing some aspects of Health-related quality of life (HRQoL). Once established, the “LeishCOM_LCL" was subjected to an assessment of face validity [26, 27].

### Face validity

Face and content validity of the core outcome instrument was assessed at the face-to-face meeting as well as through multiple virtual feedback engagement between stakeholders. Further validation was subsequently conducted by securing feedback from three independent consultant dermatologists who were not involved in the development of the outcome measures. These consultants adopted the outcome measure instrument to assess five LCL patients in each of their clinics (total number of patients (n) =15). As a result of their feedback, further and necessary amendments to LeishCOM_LCL and CRF were made. Although the authors appreciate that the face validity can be subjective [26], they approached this including multiple rounds of virtual stakeholder engagement to ensure that the CRF measured what it was intended to measure [27].

Further validation of LeishCOM_LCL was conducted through a subsequent longitudinal pilot study between March 2018 to March 2019 in a small cohort of 40 confirmed (parasitologically positive) LCL patients attending a dermatology clinic in Sri Lanka. Each patient was followed up, for a period of up to 6 months to validate the LeishCOM_LCL tool. The methodology and positive results from this further study will be published elsewhere.

## Results

### Approach to the study

We used the HOME methodology to identify core outcome domains for the development of LeishCOM_LCL as summarized in Fig 1. Following the development and judgment processes assessment of “Clinical Cure” was recognized as the overarching “General Concept”. The Core Outcome Domains identified during the development and judgment process are given in Table 1.

**Fig 1.**
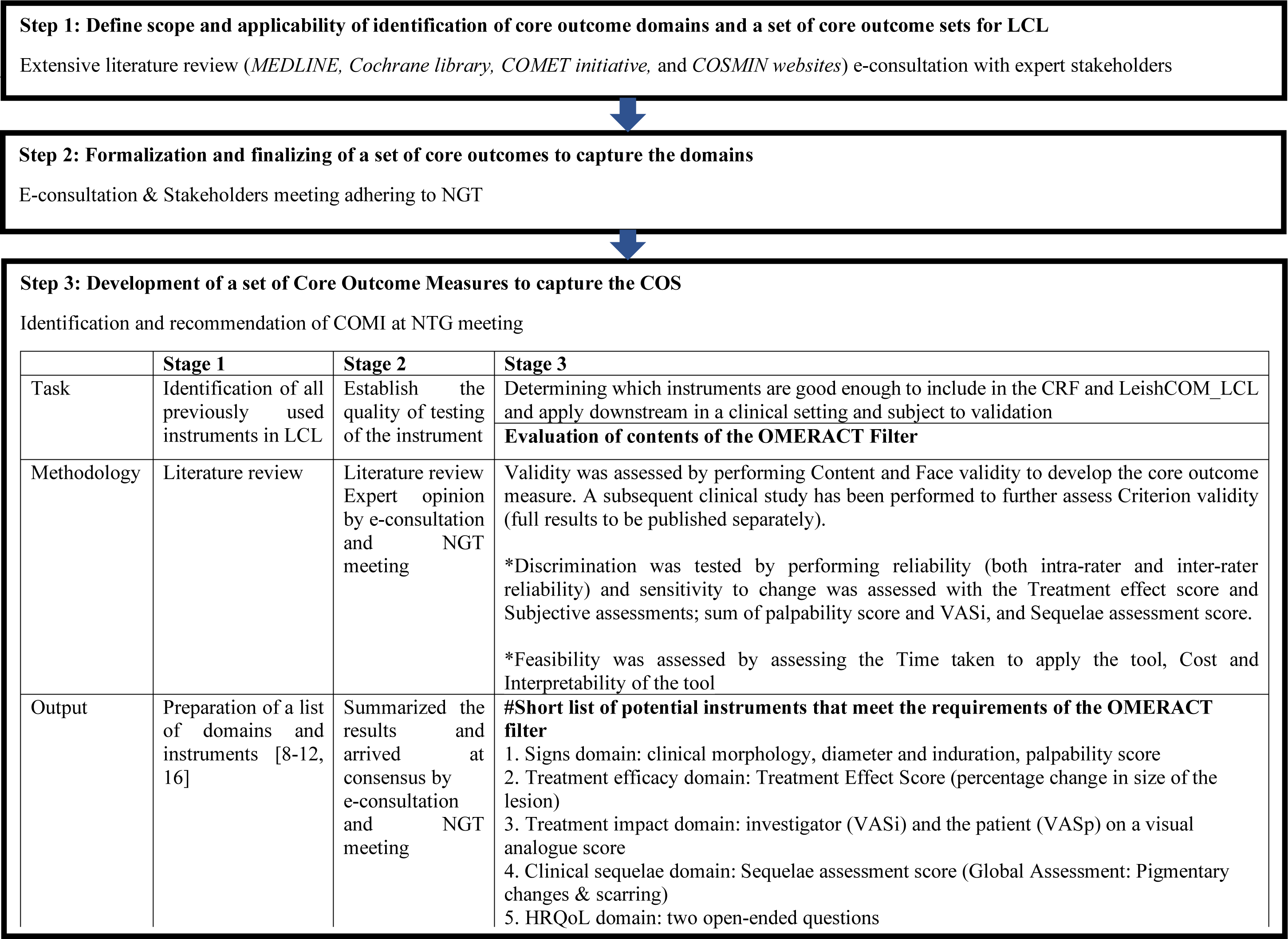
Summary of development stage of core set of outcomes in LeishCOM_LCL to each Core Outcome Domains. Stage 1-3 in accord with HOME roadmap are described. *Assessed in the Validation process. #described in detail in results section. COMI: Core outcome measure instrument, CRF: case report form, HRQoL: Health Related Quality of Life, LCL: Localized cutaneous leishmaniasis, NGT: nominal group technique, VASi: Visual analogue score investigator, VASp: Visual analogue score patient.

**Table 1.** The core outcome domains were established based on consensus.

|  | Identified Core Outcome Domains |
| --- | --- |
| 1 | Signs (Objective and Subjective assessments for localized disease/selected lesions) |
| 2 | Treatment efficacy |
| 3 | Treatment impact |
| 4 | Clinical sequelae (scarring & pigment) assessment |
| 5 | HRQoL |

### Measuring the core outcome domains

As previously described, a review of previous instruments or approaches to measure the domains was thoroughly considered. This identified that there was a paucity of measures used for LCL assessment. Therefore, each domain was discussed in detail and a standardized approach to assessing each domain was agreed on the understanding that this may require further development in the future.

#### 1. Signs

In the signs domain, to assess the primary efficacy endpoint; re-epithelialization (surface area of the ulcer) and induration, the methods described by Olliaro et al. 2013 [16] were adopted to assess the flattening of the elevated edge of the ulcerated lesions. Furthermore, a new palpability score was developed to assess both the ulcerated and non-ulcerated lesions and included in the assessment. Since erythema was not appreciated as a reliable or reproducible sign in skin of colour and pain was not considered as a universal symptom of LCL by the stakeholders, there was agreement not to measure erythema and pain. However, a free text space was provided in the CRF to record any additional signs and symptoms not captured by the agreed assessment.

#### 2. Treatment efficacy

Reduction of lesion size (re-epithelialization in an ulcerated lesion) and reduced palpability defined as flattening / reduced induration of lesions were agreed as the parameters to measure the “Treatment efficacy” domain. Erythema was not considered a reliable measure of treatment efficacy and scars and pigmentation were noted to be important sequelae which were considered in a separate domain. Table 2 outlines the clinical features for assessment as efficacy measures that were agreed through consensus at the NGT face-to-face meeting.

**Table 2:** Consensus about the important primary clinical efficacy measures were:

| Primary clinical efficacy measure | Consensus arrived at |
| --- | --- |
| Re-epithelialization for ulcerated lesions | ulcer surface area should be the primary efficacy endpoint whenever possible. |
| Flattening of non-ulcerated lesions | for non-ulcerated lesions, area of induration should be used to measure treatment efficacy |
| Absence of induration | is a valuable efficacy measure but acknowledged as difficult to standardize |
| Overall erythema | was thought **not** sufficiently reliable to act as a measure of treatment efficacy especially in skin of colour. |
| Presence of scars and pigmentation | were recognized as important sequelae that required assessment/grading as part of a separate domain. |

#### 3. Treatment Impact

The treatment impact domain was recorded on each day of assessment by capturing the perception by both the investigator and the patient on a visual analogue score.

#### 4. Clinical Sequelae

Pigmentary change (Hypo/hyper) and scarring (atrophic or hypertrophic) were recognized as the parameters to capture for the clinical sequelae assessment domain. It was noted that either hypo or hyperpigmentation could result from LCL and scars could be either atrophic or hypertrophic. Therefore, all 4 changes were considered as clinical areas suitable for rating.

#### 5. HRQoL

The impact of the HRQoL had not been previously considered in LCL and therefore two open-ended questions were included as a preliminary step to try and capture the most important aspects for the patient with a view to informing a novel tool at a later date. It was decided to include open-ended questions to make it easy and straightforward for the patient to respond and to capture the most important aspect of thoughts originated by the patient. This would further ensure that clinicians recognize patient’s problems and consider these in patient management.

### Generation of the Case Report Form (S Appendix 1)

A Case Report Form (CRF) was generated to document the demographic details and reflect the agreed COS (Core Outcome Domains and Measurements) in each patient. The CRF contained a cover page noting the document category, code, title of the CRF, approved version, sponsor, date of release, authorization from the Principal Investigator, and a table of contents to guide the user. The next two pages of the CRF contained instructions for the user on individual items. Written as well as diagrammatic and photographic instructions with clinical examples were provided for measuring and assessing LCL to minimize any potential ambiguities. The rest of the pages contained demographic details of the patient, enrollment particulars; details of obtaining consent, slit skin smear and or punch biopsy details, relevant clinical history and examination details, assessments at baseline, 4 weeks, 3 months, and 6 months from the onset of treatment, a summary of scores over time, details on drug therapy, selected investigation results with dates, final comments, and the investigator’s signature with the date. The day of enrollment into the study was taken as the “Baseline”. Each time point was calculated from the Baseline. After several further rounds of feedback and revisions from experts as described in Methods, the 14^th^ Version of the CRF with clinical score was agreed as the final consensus version (S Appendix 1).

### Capture of Signs domain (Objective and Subjective assessments)

#### a) Guidelines for selection of lesion to follow up and validation of OMIs

In the context of the study, as patients may have more than one lesion of LCL, guidelines were provided in the CRF to select an “index lesion” to be used throughout the period of clinical assessment. The index lesion represented a recent onset, clinically typical looking localised CL lesion which was confirmed with positive parasitology. Each type of lesion was well described (Supplementary Table 1). There was opportunity for the investigator to report “Any other atypical lesions” on the CRF during examination (CRF Section 5.1.14). A further Section 5.1.15 in the CRF captured “Patient reported symptoms e.g. pain, loss of function etc.” This was to build up in the future if any useful signs were reported by the patient during the validation process. The anatomical location of the lesion was identified on a body diagram. A space was provided to record the biopsy site if taken. It was made compulsory to have a laboratory confirmed diagnosis (either the presence of *Leishmania* amastigote in a slit skin smear/biopsy and histology or positive PCR) to enroll patients during the downstream application of CRF on patients in the validation process of LeishCOM_LCL.

##### Outcome measurement instrument for signs

Objective assessment (lesion measurements with a ruler & ball-point pen) (S Fig 1 & 2) was described for ulcerated lesions as recommended by the previous harmonised guidance paper [16]. Both size of the ulcer and palpability of the induration were taken into consideration. As the panel perceived that palpability was an important feature of disease activity, a newly developed subjective assessment (a palpability score of 0,3,6,9) was described for both non-ulcerated and ulcerated lesions (0=flat, 9=severly raised) (Tables 3 & 4). Schematic images and/or photos alongside descriptions were used to standardise the assessment.

**Table 3.**
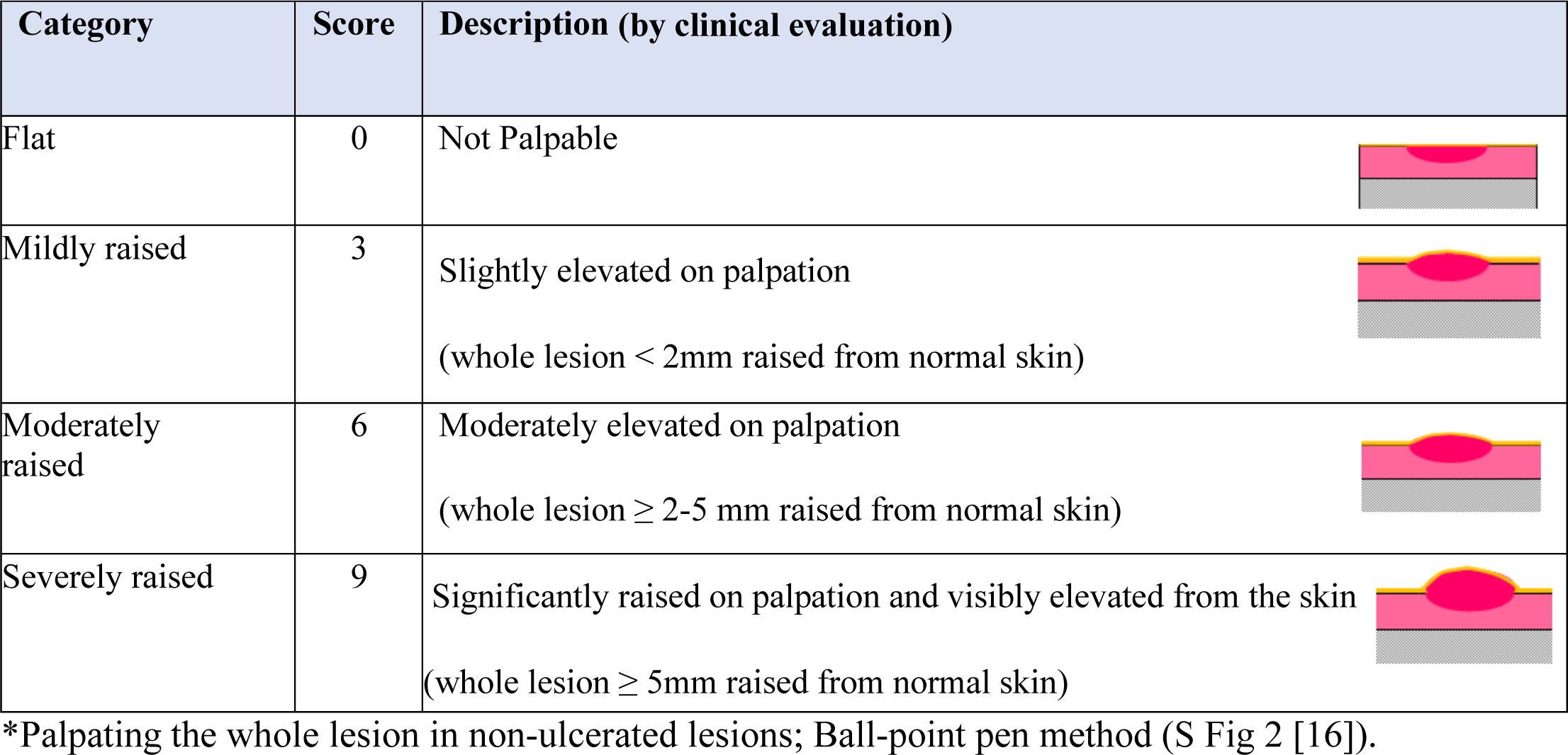
Description of the palpability score for non-ulcerated lesions*.

| Category | Score | Description (by clinical evaluation) |
| --- | --- | --- |
| Flat | 0 | Not Palpable |
| Mildly raised | 3 | Slightly elevated on palpation<br>(whole lesion < 2mm raised from normal skin) |
| Moderately raised | 6 | Moderately elevated on palpation<br>(whole lesion ≥ 2-5 mm raised from normal skin) |
| Severely raised | 9 | Significantly raised on palpation and visibly elevated from the skin<br>(whole lesion ≥ 5mm raised from normal skin) |

\*Palpating the whole lesion in non-ulcerated lesions; Ball-point pen method (S Fig 2 [16]).

**Table 4.** Description of the palpability score based on the edge for ulcerated lesions*.

| Category | Score | Description (by clinical evaluation) |
| --- | --- | --- |
| Flat | 0 | Not Palpable |
| Mildly raised | 3 | Slightly elevated on palpation<br>(edge of the lesion < 2mm raised from normal skin) |
| Moderately raised | 6 | Moderately elevated on palpation<br>(edge of the lesion ≥ 2-5 mm raised from normal skin) |
| Severely raised | 9 | Significantly raised on palpation and visibly elevated from the skin.<br>(edge of the lesion ≥ 5 mm raised from normal skin) |

\* Instructions: for ulcerated lesions measure by palpating the “edge” of the ulcerated lesion.

### Capture of “Treament Efficacy” domain

#### Treatment effect score

“Treatment Effect Score” was assessed at week 4 and at 3 & 6 months from initiation of treatment. Since there are no specific guidelines on designing numerical scores, and different numerical scores have been used successfully to predict clinical outcomes [28], the clinicians present at the NGT meeting agreed to rate the “Treatment Efficacy” with scores of 12, 9, 6, 3, 0; a score of “12” was rated for no improvement and “0” for complete clinical cure. The Treatment Effect Score mainly took into account the percentage change of the lesion size from baseline but also embraced factors about re-epithelialization and inflammation at each assessment point in comparison with baseline as a means of trying to prevent any ambiguity and ensure some consistency between raters (Table 5). Although erythema was considered as "**not** sufficiently reliable to act as a measure of treatment efficacy especially in skin of colour", it was decided to include an assessment of "inflammation” (using subjective assessment of erythema by clinical-eyeballing) when doing an Investigator Global assessment of the overall Treatment Effect Score as acute inflammation is known to subside/disappear with wound healing [29]. This was combined with other anticipated features expected with therapeutic resolution of a lesion.

**Table 5.** Treatment effect score: Description of Investigator Global Assessment of an active disease post-treatment.

| Score | Expected features | Allocate Score |
| --- | --- | --- |
| 12 | No improvement. Lesion remained active, having the same characteristics, or becoming larger than prior to the **start of treatment**. |  |
| 9 | Size of the lesion decreased 50% in comparison with the initial lesion, with fewer inflammatory signs* and discrete re-epithelialization (Size: diameter, length & width) |  |
| 6 | Size of the lesion decreased between 50–90% in comparison with the initial lesion, and left few inflammatory signs* |  |
| 3 | Size of the lesion decreased more than 90%, with re-epithelialization and very little inflammation* |  |
| 0 | Complete re-epithelialization with a characteristic scar and no inflammation. Active disease settled |  |

\*Inflammatory signs: erythema by clinical-eyeballing and having anticipated features expected with therapeutic resolution of a lesion

### Capture of “Treatment Impact” Domain

#### Treatment impact score

##### Visual Analogue Score (VAS)

To measure the impact of the treatment on the “skin problem” at the time of assessment a Visual Analogue Score (VAS) ranging from 0 - 10 (0=“completely clear skin”, 10=“severely affected skin”) was described for both investigator (VASi) and patient (VASp) (CRF Section 6.3). Patients and investigators were asked to consider how they would score the skin problem on the day of assessment starting at baseline and after commencing treatment. Options were provided for the investigator and the patient to put a mark on the line to indicate how adversely they perceived the skin was affected on the day of the assessment (at Baseline, 4 weeks, 3 months & 6 months from the onset of treatment). The line of a VAS is 10cm in length and a score is allocated according to the nearest whole cm (Fig 2).

**Fig 2.**
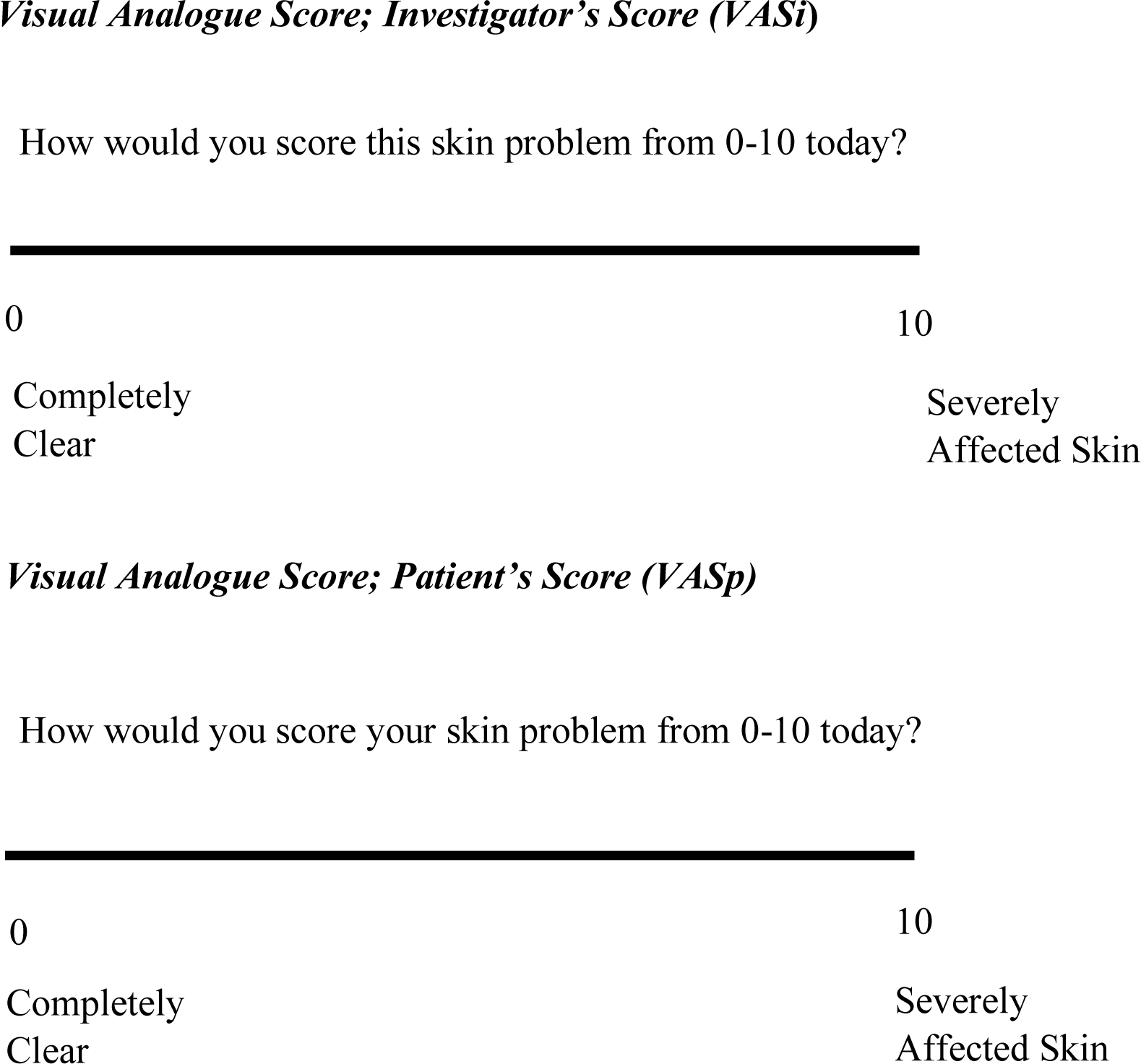
Visual analogue score of the investigator (VASi) and patient (VASp). Each line was 10 cm long. Each score is to be allocated according to the nearest whole cm.

#### Capture of Clinical sequelae (scarring & pigment) assessment domain

As sequelae including scarring and pigment changes are a common occurrence from LCL, a new Investigator Global Sequelae Assessment score was developed which consisted of “pigment change, atrophic scars, and hypertrophic/keloid scars”. Each of these items was rated from 0-3. The aim was to establish the frequency of development of sequelae and also to try and assess whether earlier effective therapy might reduce the likelihood of sequelae. The investigator is asked to allocate a subjective score to the sequelae assessment. The scoring of pigmentation and scarring was discussed in detail at the NGT judgment process, and it was decided to compare the colour change in the lesion and the surrounding area of the lesion with the opposite unaffected side of the body. Furthermore, photographs taken at the field visit were examined in detail at the NTG meeting and scores ranging from 0-3 were allocated for pigment change by consensus (Table 6, and S Fig 3 & 4). Scarring was decided to be assessed by palpation and by close clinical examination (Table 6, and S Fig 5 & 6).

**Table 6.** Investigator Global Assessment of i & ii) Pigment change iii) Atrophic scars iv) Hypertrophic/ Keloid scars.

| Score (0-3) Pigment Change (Hyperpigmentation) |  |  | Allocate Score |
| --- | --- | --- | --- |
| Category | Score | Description |  |
|  | 0 | No hyperpigmentation |  |
|  | 1 | Mild hyperpigmentation |  |
|  | 2 | Moderate hyperpigmentation |  |
|  | 3 | Severe hyperpigmentation |  |
| Score (0-3) Pigment Change (Hypopigmentation) |  |  |  |
| Category | Score | Description |  |
|  | 0 | No hypopigmentation |  |
|  | 1 | Mild hypopigmentation |  |
|  | 2 | Moderate hypopigmentation |  |
|  | 3 | Severe hypopigmentation |  |
| Score (0-3) Atrophic scars |  |  |  |
| Category | Score | Description |  |
| Clear | 0 | No scar visible or detectable on palpation |  |
| Mild | 1 | Minimal atrophic scarring – little change on palpation |  |
| Moderate | 2 | Atrophic scarring with textural changes of skin |  |
| Severe | 3 | Deep atrophic / mutilating scar |  |

| **Score (0-3) Hypertrophic / Keloid scars** |  |  |
| --- | --- | --- |
| **Category** | **Score** | **Description** |
| Clear | 0 | No scar visible or detectable on palpation |
| Mild | 1 | Minimal hypertrophic scarring - some palpable change |
| Moderate | 2 | Palpable scarring with textural changes of the skin |
| Severe | 3 | Mutilating scar (with underlying structural involvement) |
| **TOTAL SCORE** |  |  |

### Summarizing the scores of the LeishCOM_LCL

A table to summarize the subjective scores (palpability score & VASi & VASp) and sequelae assessment scores were included at the end of the clinical score (Table 7). The summary of scores is to be calculated by each investigator/rater at the end of the assessment at each time point and entered in the table (Table 7). These data will be used later for analysis and to arrive at conclusions during clinical trials or at routine treatment clinics (manuscript is being prepared in the completed validation stage).

**Table 7.** Summary of scores.

| **SUBJECTIVE** | **SCORE** |
| --- | --- |
| Palpability; Non-ulcerated lesions | (0-9) |
| Palpability; Ulcerated lesions | (0-9) |
| Visual Analogue; Investigator | (0-10) |
| Visual Analogue; Patient | (0-10) |
| **TOTAL** |  |
| **TREATMENT EFFECT** | **SCORE** |
| Investigator Assessment | (0-12) |
| **TOTAL** |  |
| **SEQUELAE ASSESSMENTS** | **SCORE** |
| Pigment Change; Hyperpigmentation | (0-3) |
| Pigment Change; Hypopigmentation | (0-3) |
| Atrophic Scars | (0-3) |
| Hypertrophic/Keloid Scars | (0-3) |
| **TOTAL** |  |

#### Capture of Health-Related Quality of Life (HRQoL) domains

Studies examining HRQoL in LCL are limited and the published studies have not necessarily acknowledged negative impacts including those caused by treatment [30, 31]. Thus our OMI incorporated two open-ended questions “How does your skin problem affect you?” and “What are the three worst aspects of having your skin problem?” The aim was to assess the patient’s perspectives with a view to adapting the tool or developing a relevant patient reported outcome measure encompassing HRQoL assessment in the future. Information on the two open-ended questions was gathered from patients and thematic analysis was carried out during the validation process in the downstream application of the CRF in a dermatology clinic in Sri Lanka. Psychological impact improved in line with treatment response over a 6 month period from baseline, however, 30% of patients expressed psychological concerns as a result of sequelae such as pigment changes and scarring (Full data and results will be published in a separate manuscript).

### Face Validity

The face validity was established on parameters regarding appropriateness of grammar, clarity and unambiguity of items, correct spelling of words, correct structuring of sentences, appropriateness, and adequacy of instruction on the instrument, structure of the instrument in terms of construction and, appropriateness of difficulty level of the instrument for the participants, and reasonableness of items in relation to the purpose of the instrument [27]. The content was addressed following feedback sent by the three consultant dermatologists by applying the OMI to five patients at each of their clinics (total patient number (n=15)) were considered before finalizing and revisions were made by experts with 100% agreement.

Further positive results of validity testing of the LeishCOM_LCL has been established including criterion validity. The robust methodology and results from this will be published separately.

## Discussion

This paper includes detailed development of a COS for LCL with identification of Core Outcome Domains, measurement of the domains and a development of a subsequent OMI through adoption of stages 1, 2 & 3 of the HOME roadmap [19, 24]. This study was initiated as in-depth literature review revealed the absence of a standardized and validated COS for LCL to assess response to current and novel treatment measures in clinical trials and clinical practice across the globe. This scarcity has led to an overall inability of comparison between trials and recommendations of best of care of management for LCL patients [8]. There was only one study that described a few harmonized outcome measures for use in LCL clinical trials [16] which informed our LeishCOM_LCL. Our study is the first to identify a set of Core Outcome Domains for LCL using recognised and robust methodology in a standardized manner. The study has also included consideration of how to measure the agreed domains in LCL as a means of developing a core outcome set for use in the assessment of LCL. These have informed an outcome measurement tool for LCL (LeishCOM_LCL). As no outcome measures have previously been agreed by broad consensus for each domain, our group has developed and suggested an approach for each domain and incorporated these measures into a practical tool LeishCOM_LCL.

The NGT adopted during the development stage of this study is a valid technique, representing an alternative to the Delphi methodology [24]. Participation at the NGT meeting provided opportunity for open dialogue with a moderator and provided time for clinical presentations and translation where necessary.

This novel tool captures and scores relevant objective and subjective clinical outcomes embracing active signs including sequelae and takes into account the perspectives of both the patient’s and investigator’s with respect to the healing process and treatment of LCL lesions. A visual analogue score was used to capture both participants’ and investigators’ perspectives as VAS is known to be a valid, reliable, and repeatable method of assessment of therapeutic response in other dermatological diseases [32]. The validation process with the small cohort of patients showed that this tool LeishCOM_LCL is reliable with a good face and content validity. This tool will also be useful to assess cure rates, treatment failure, relapse rate and to assess the other case management indicators described in the Manual for case management of cutaneous leishmaniasis in the WHO Eastern Mediterranean Region [17]. Further criterion validity has been performed in a larger cohort and the robust methodology and positive results secured will be published in a separate manuscript.

In recent years HRQoL and patient-reported outcome (PROs) have been considered as a very important part of ensuring a patient-centered approach in disease management [33]. Addressing HRQoL in a systematic manner was beyond the scope of this study. However, we recognize that additional measures could further enhance the assessment of LCL particularly in respect of capturing patient-reported outcomes and HRQoL. In LeishCOM_LCL, two open-ended HRQoL questions “How does your skin problem affect you?” and “what are the worst aspects of having your skin problem?” were used to capture patients’ perceptions of having LCL and the issues they face during prolonged treatment. We adopted this pragmatic approach in the first instance to try and ensure the patient’s perspective was recorded on paper. No previous study to date has attempted to record/report these aspects. The results from this approach were analyzed during the validation process and this highlighted the need to ensure adverse effects from treatments and negative impacts of LCL are fully recognized when assessing this disease thus enabling a patient-focused and empathic approach to management (details are due for publication in a validation paper). The authors suggest that this work could help to inform the development of a more robust PROM specific to LCL in the future and the authors appreciate that systematic qualitative research with audio recordings would be helpful to expand upon this area with robust analysis.

The lack of a more diverse group of stakeholders including dermatologists from the Mediterranean region and Africa, patients from diverse geographic areas, and pharmaceutical industry representation was another limitation in this study. However, the stakeholders involved represented important endemic regions for LCL and the authors acknowledge that further improvement and fine-tuning of this OMI may be achieved by including further stakeholders from other LCL endemic regions with a global representation and further individual outcome measures may need to be developed for each Core Outcome Domain, particularly relating to HRQoL. The team also appreciate further engagement with patients from each region as well as personnel from the pharmaceutical industries and regulatory bodies could inform future discussions and adoption. Furthermore, it will be important in the future to assess whether, scarring and pigmentation should remain a primary efficacy endpoint or secondary efficacy endpoint and how these might impact HRQoL. Data analysis secured from the validation process will help to inform future improvements and this approach will complete stage 4 & 5 of HOME methodology.

The quality assurance stage (Stages 4 & 5); validity, reliability, responsiveness, interpretability and feasibility of scoring and HRQoL had already been assessed in the newly developed LeishCOM_LCL tool by applying the OMI downstream in a dermatology clinic to a small cohort of patients (n=40) in Sri Lanka in accord with the HOME roadmap [24], COSMIN [34] and Guidance for Industry Patient-Reported Outcome Measures: Use in Medical Product Development to Support Labeling Claims, U.S. Department of Health and Human Services Food and Drug Administration Center 2009 [35]. This will be presented in a future manuscript.

In conclusion, the Core Outcome Domains i.e. what to measure in LCL have now been defined through a process of consensus. Agreement on how to measure the agreed Core Outcome Domains was secured following literature review and a multidisciplinary and international stakeholder meeting. The LeishCOM_LCL is the first OMI to be developed in a standardized manner to assess LCL and therefore provides potential for broad adoption for use in clinical trials and routine clinical settings.

Our future aim is to update the LeishCOM_LCL to reflect important views of patients when collecting information and to consider developing a PROM specific to LCL. A specific PROM should ensure adverse effects from treatments as well as the negative impacts of LCL are captured when assessing this disease thus enabling a patient-focused and empathic approach to management.

## Data Availability

All data produced in the present work are contained in the manuscript

## Acknowledgments

We thank and acknowledge the Sri Lanka College of Dermatologists for giving their immense support during the development and validation process of this OMI

## Supplementary tables

**S1 Table.** Guidelines for clinical categorization of the Index Lesion*.

|  |
| --- |
| **Presenting sign(s): Please indicate which of the following clinical features are evident and the number of lesions at the time of presentation and if possible, the duration of the lesion (s).** |
| 5.1.1 Recent onset macule (circumscribed change in the color of skin that is flat on palpation – (excludes scarring and post inflammatory pigmentary change) |
| 5.1.2 Papule ($\leq 5$mm diameter, palpable solid elevation) |
| 5.1.3 Nodule ($> 5$ mm diameter, palpable elevation) |
| 5.1.4 Plaque (flat topped with diameter greater than its height) |
| **Ulcerative change** |
| 5.1.5 Dry ulcer (destruction of epidermis of skin with central crusting/scaling) |
| 5.1.6 Wet ulcer (destruction of epidermis of skin with wet exudates) |
| 5.1.7 Nodular ulcerative ($> 5$mm diameter, palpable elevation with central ulceration) |
| **Other features associated with acute lesion(s)** |
| 5.1.8 Satellite lesions |
| 5.1.9 Halo pigmentation |

\*An active lesion of most recent onset which was parasitologically confirmed has to be selected
as an “**index lesion**” to be assessed throughout the study from one time point to another.

## Supplementary Figures

**S Fig 1.**
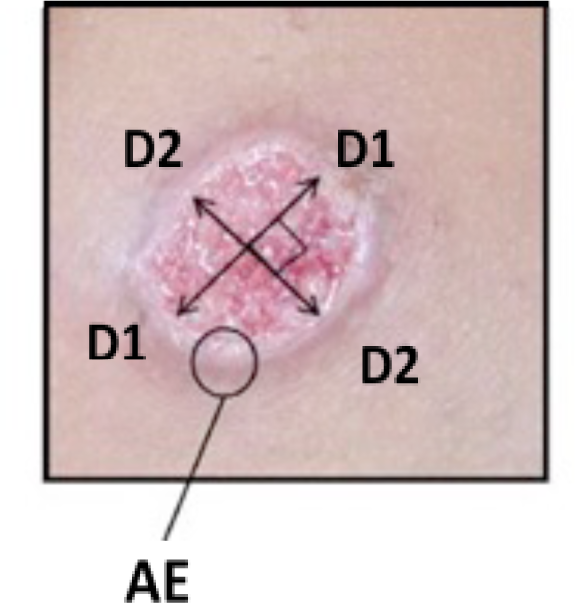
Instructions to measure the diameters of an ulcerated lesion. Measure the largest diameter of the ulcerated area [D1] and then select the largest diameter that is perpendicular to the original measurement taken [D2]. If adherent crust evident, assess the 2 largest diameters of the crusted area in the same way [8]. AE: Elevated active edge of the lesion.

**S Fig 2.**
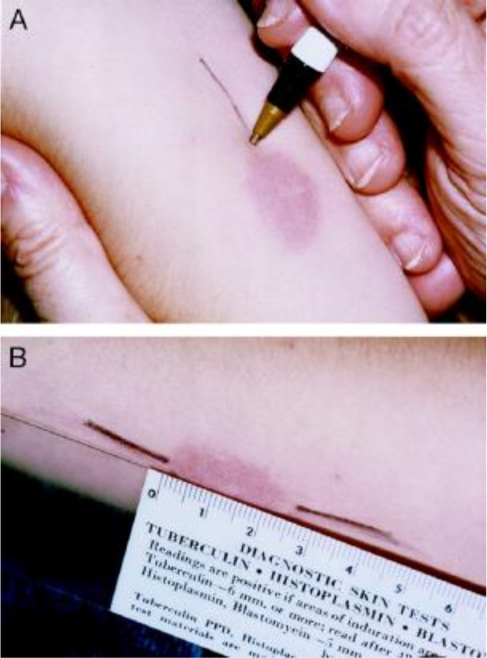
Instructions to measure the diameters of the indurated area of a non-ulcerated lesion. Identify the widest diameter of the lesion and draw two lines up to the edge of the lesion in line with the largest diameter [A] and then measure the distance between the two lines [B]. Similarly find the largest diameter that is perpendicular to the original measurement taken as above.

**S Fig 3.**
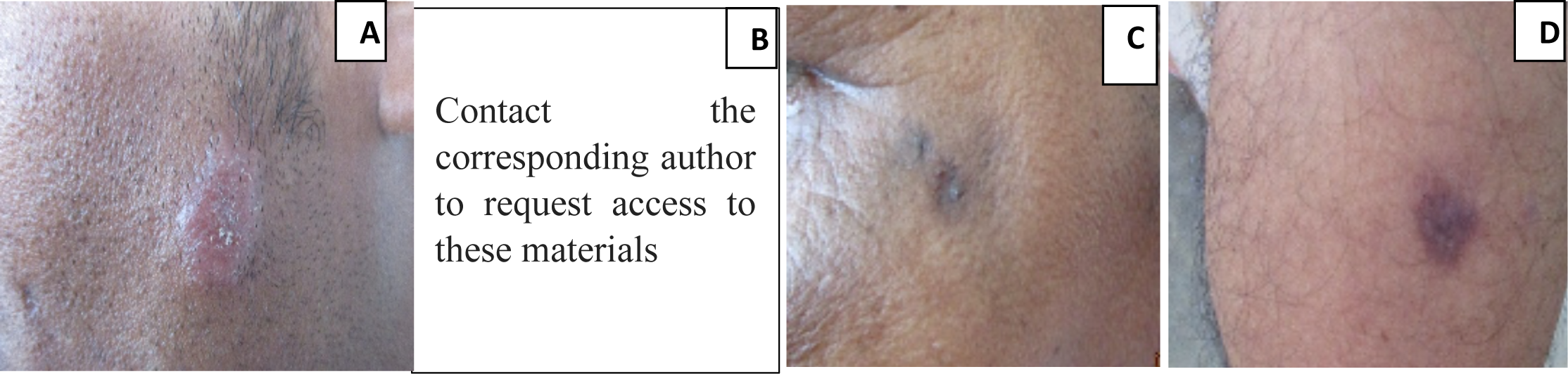
Grading of hyperpigmentation during the NGT meeting. A: no hyperpigmentation, B: mild hyperpigmentation, C: moderate hyperpigmentation, D: severe hyperpigmentation. NGT: Nominal group technique

**S Fig 4.**
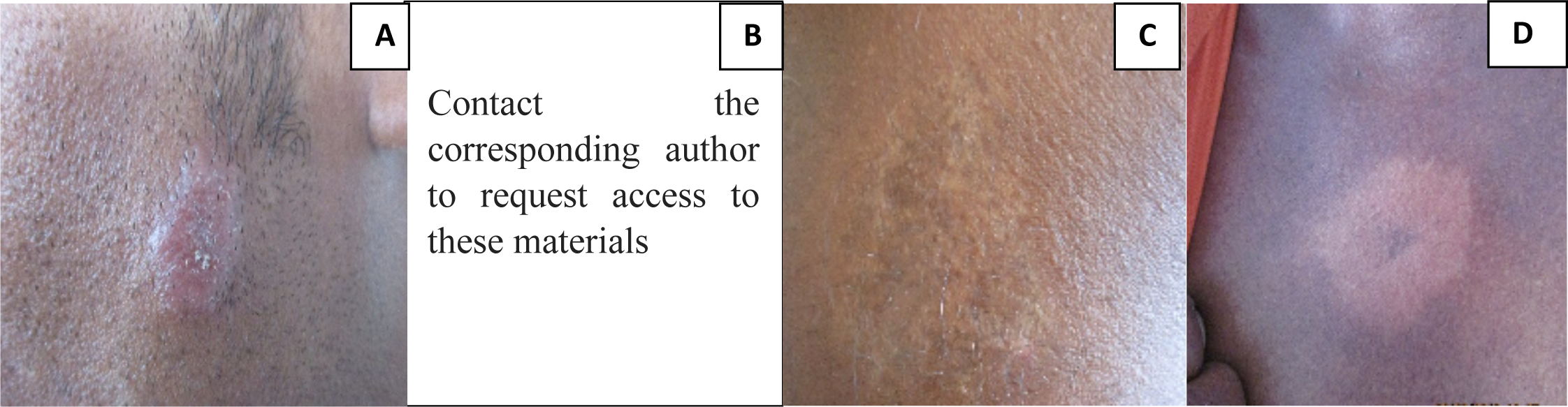
Grading of hypopigmentation during the NGT meeting. A: no hypopigmentation, B: mild hypopigmentation, C: moderate hypopigmentation, D: severe hypopigmentation.

**S Fig 5.**
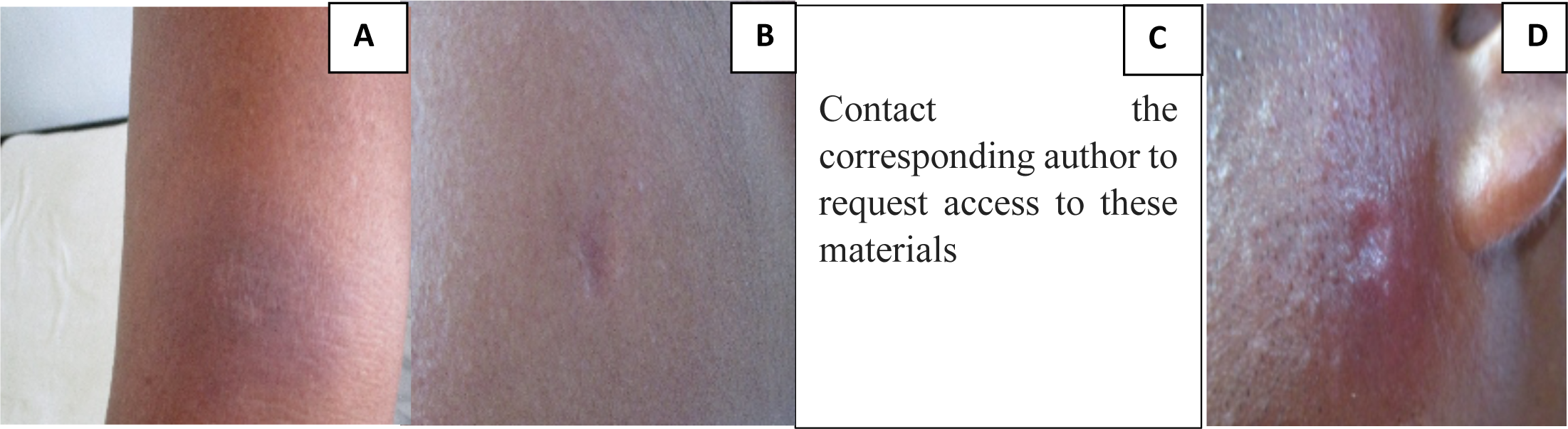
Grading of atrophic scarring during the NGT meeting. A: no atrophic scarring, B: mild atrophic scarring, C: moderate atrophic scarring, D: severe atrophic scarring.

**S Fig 6.**
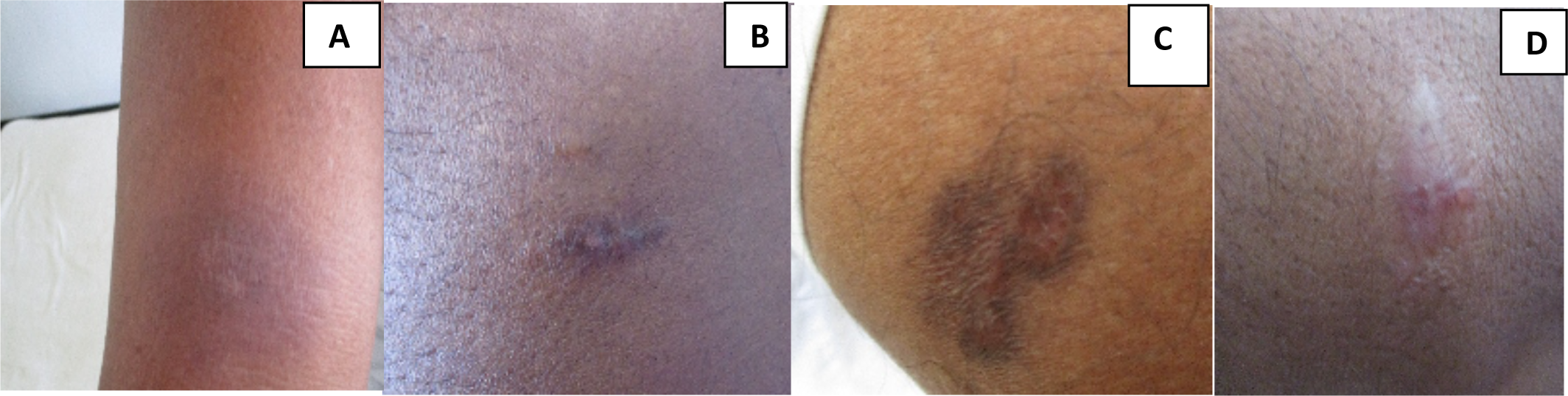
Grading of hypertrophic scarring during the NGT meeting. A: no hypertrophic scarring, B: mild hypertrophic scarring, C: moderate hypertrophic scarring, D: severe hypertrophic scarring.

## CASE REPORT FORM FOR LOCALISED CUTANEOUS LEISHMANIASIS

**Towards a global research network for the molecular pathological stratification of leishmaniasis**

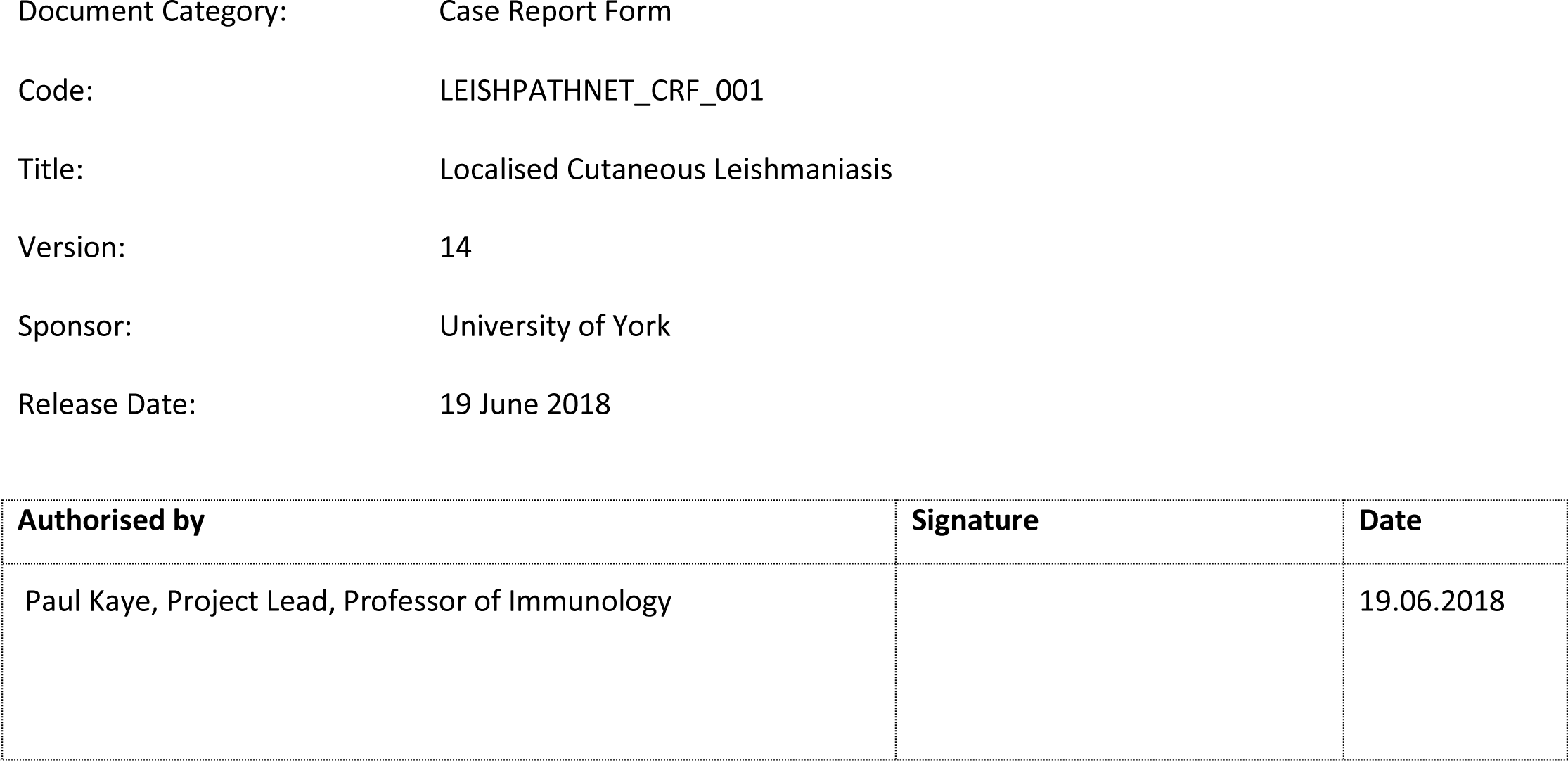

### 1. INSTRUCTIONS FOR MEASURING AND ASSESSING LOCALISED CUTANEOUS LEISHMANIASIS

#### DEFINING LOCALISED CUTANEOUS LEISHMANIASIS

Include patients with up to 5 lesions. Please take a photo of the lesions as per SOP for photography.

#### INSTRUCTIONS FOR IDENTIFYING LESIONS AND TAKING BIOPSIES

Clearly identify which lesions will be assessed at each visit and where biopsies have been taken on the figures in the CRF

Provide a description of any lesion(s) biopsied and / or being assessed at each visit in the table below the figures as indicated.

#### OBJECTIVE ASSESSMENTS FOR LOCALISED DISEASE/SELECTED LESIONS

##### a) Ulcer size

Measure the largest diameter of the ulcerated area [D1] and then select the largest diameter that is perpendicular to the original measurement taken [D2]. If adherent crust evident, assess the 2 largest diameters of the crusted area in the same way [16].

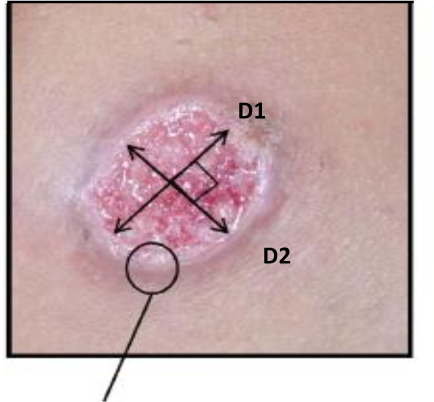

##### b) Area of induration of the lesion

Identify the widest diameter of the lesion and draw two lines up to the edge of the lesion in line with the largest diameter and then measure the distance between the two lines. Similarly find the largest diameter that is perpendicular to the original measurement taken as above.

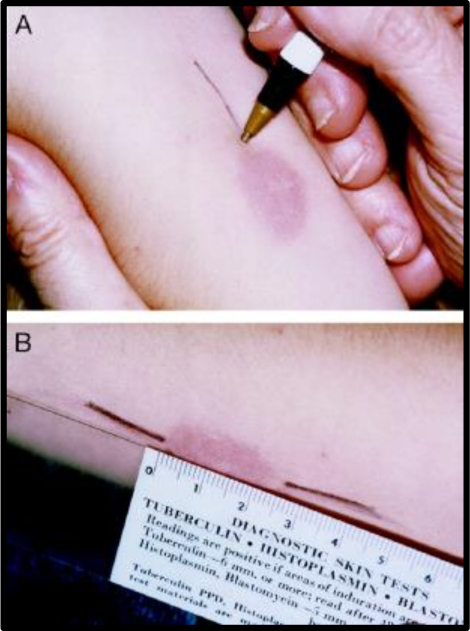

#### SUBJECTIVE ASSESSMENTS FOR LOCALISED DISEASE/SELECTED LESIONS

For **<u>non-ulcerated</u>** areas measure by palpating the whole lesion and indicate a palpability score as below

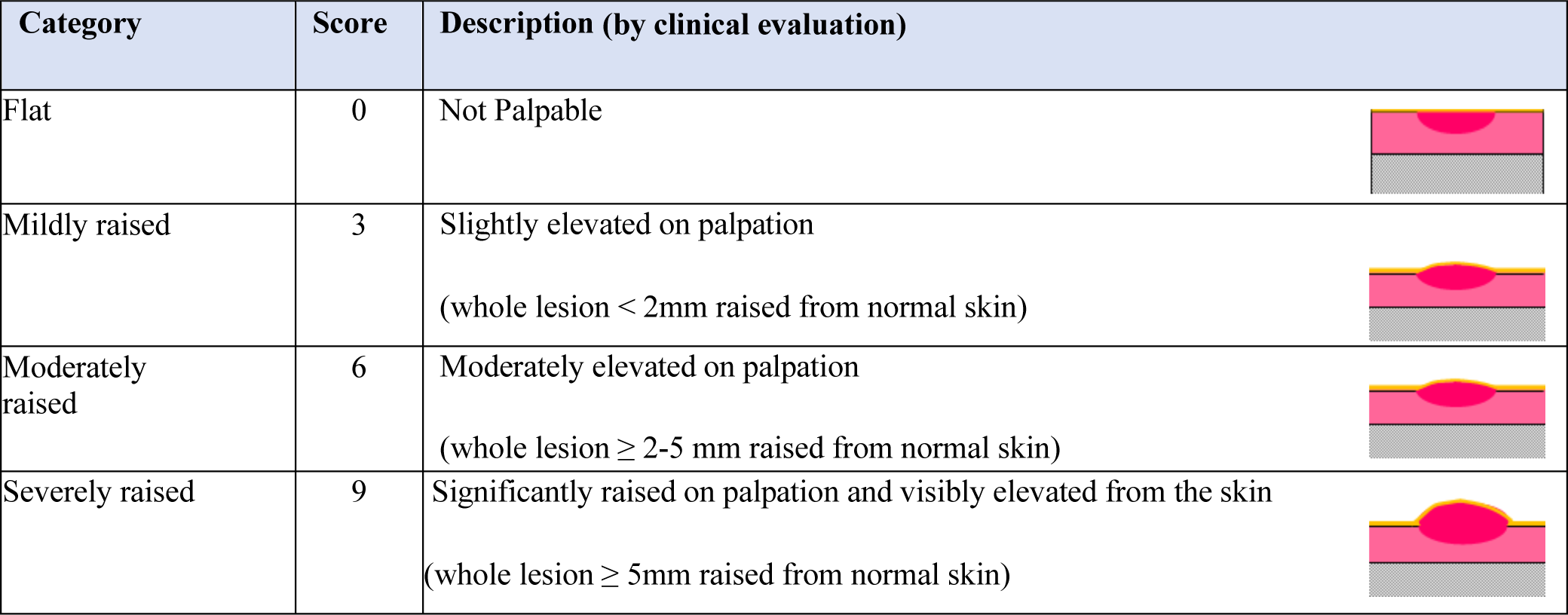

For **<u>ulcerated lesions</u>** measure by palpating the EDGE of the lesion

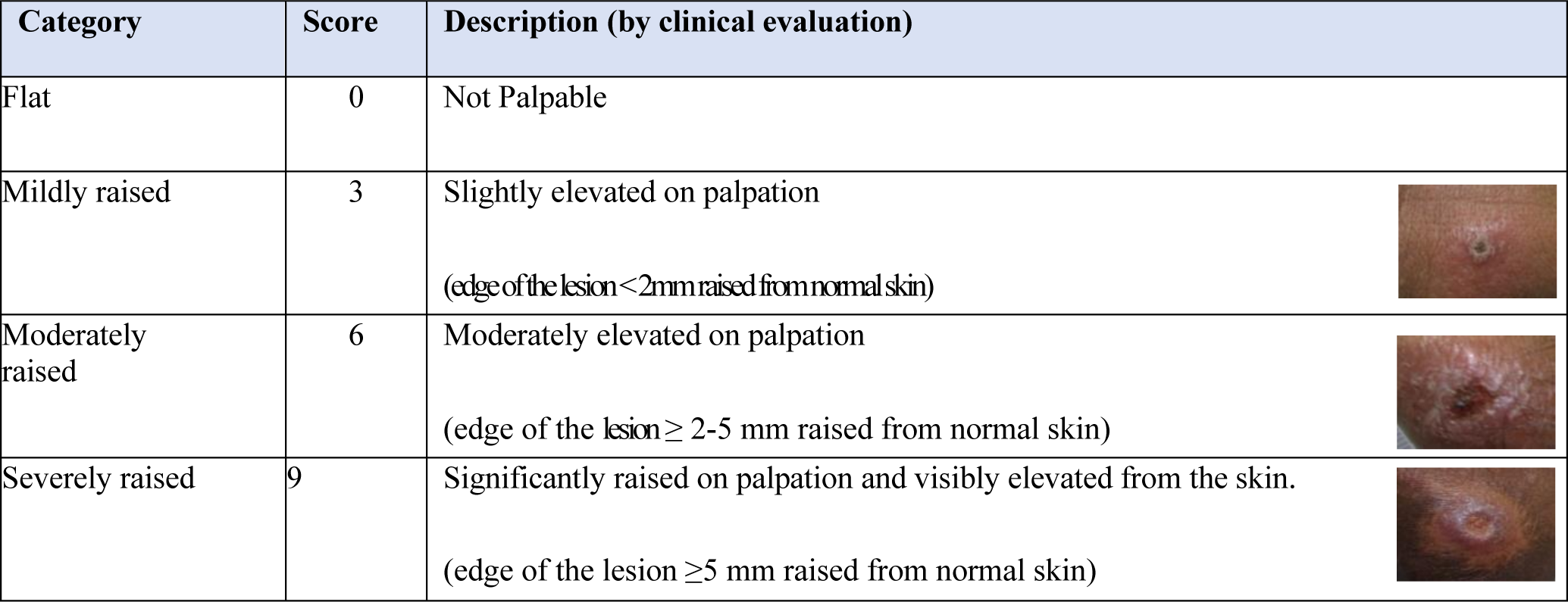

#### VISUAL ANALOGUE SCORE

Please ask healthcare professional **AND** the patient to put a mark on the line to indicate how badly they perceive the skin is affected **on the day of the assessment**. The line is 10cm and the score will be allocated according to the nearest whole cm. 0 represents clear skin and 10 represents the worse the skin can get.

#### TREATMENT EFFECT SCORES

Please indicate how much improvement there has been at visits 4 weeks, 3 months and 6 months

#### SEQUELAE ASSESSMENTS

Please score the various potential sequelae from 0-3

#### MEASURING HRQoL

Please ask patients two open ended questions, as indicated in the CRF.

#### DATA FOR CASE REPORT FORM FOR LOCALISED CUTANEOUS LEISHMANIASIS

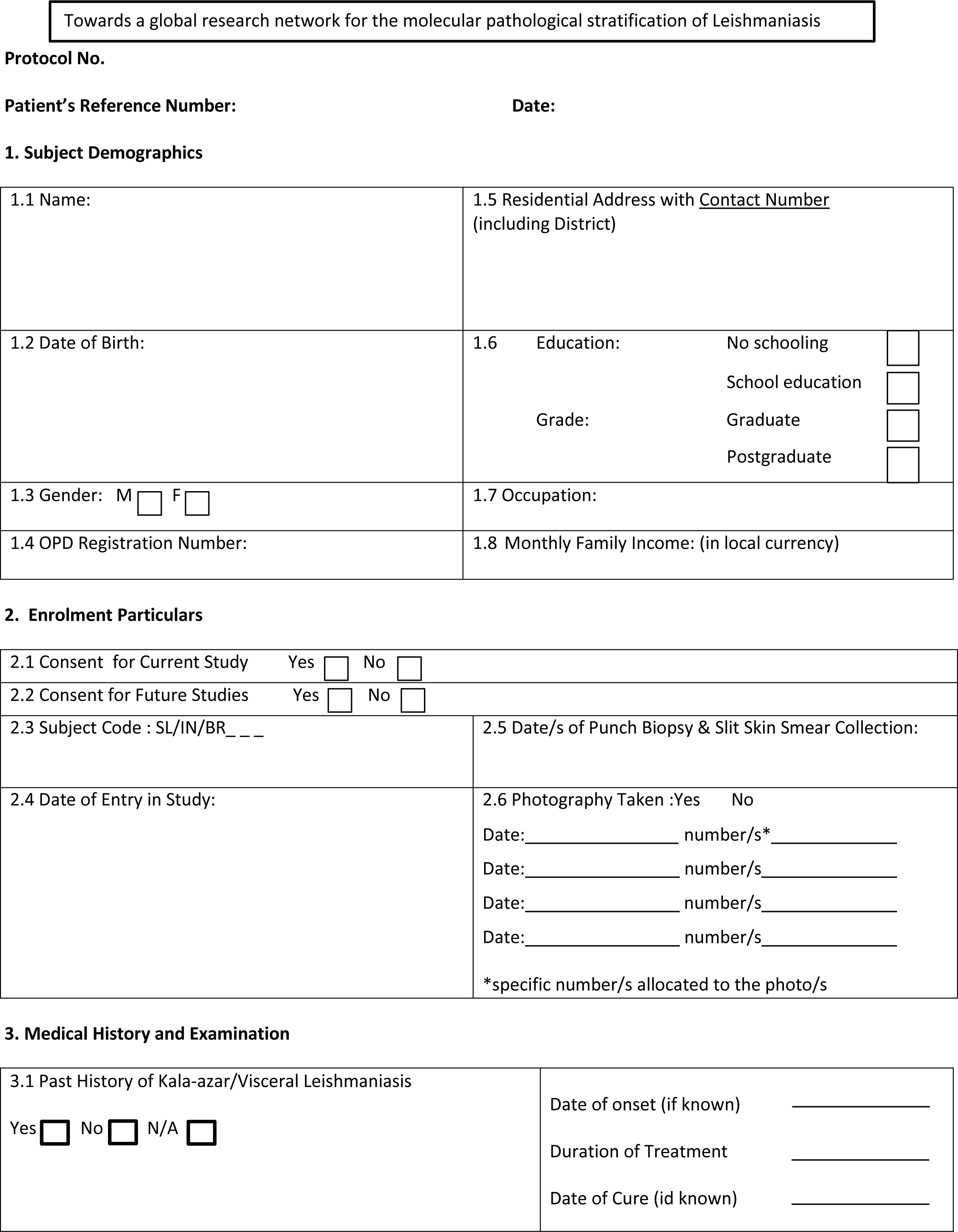
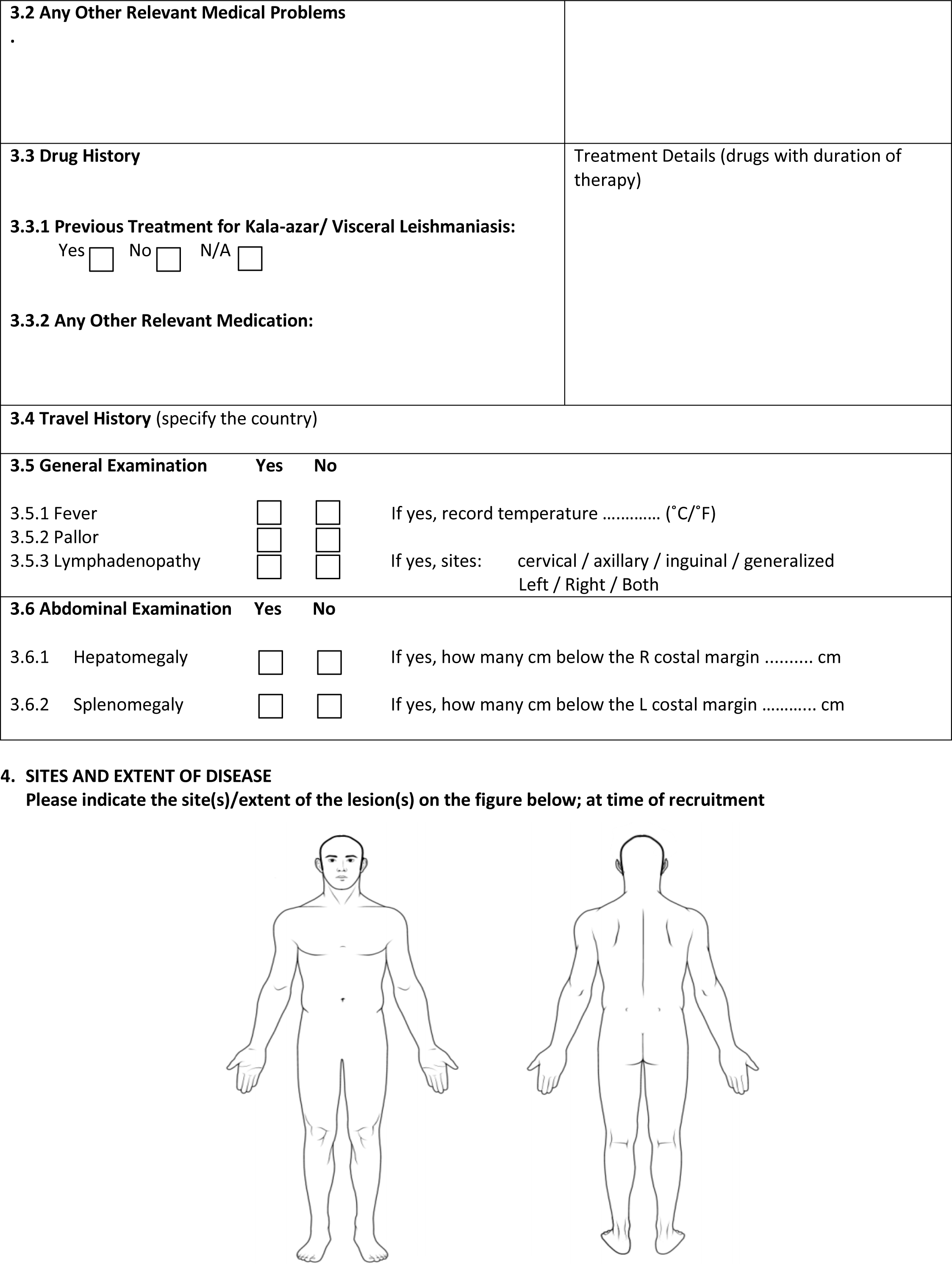
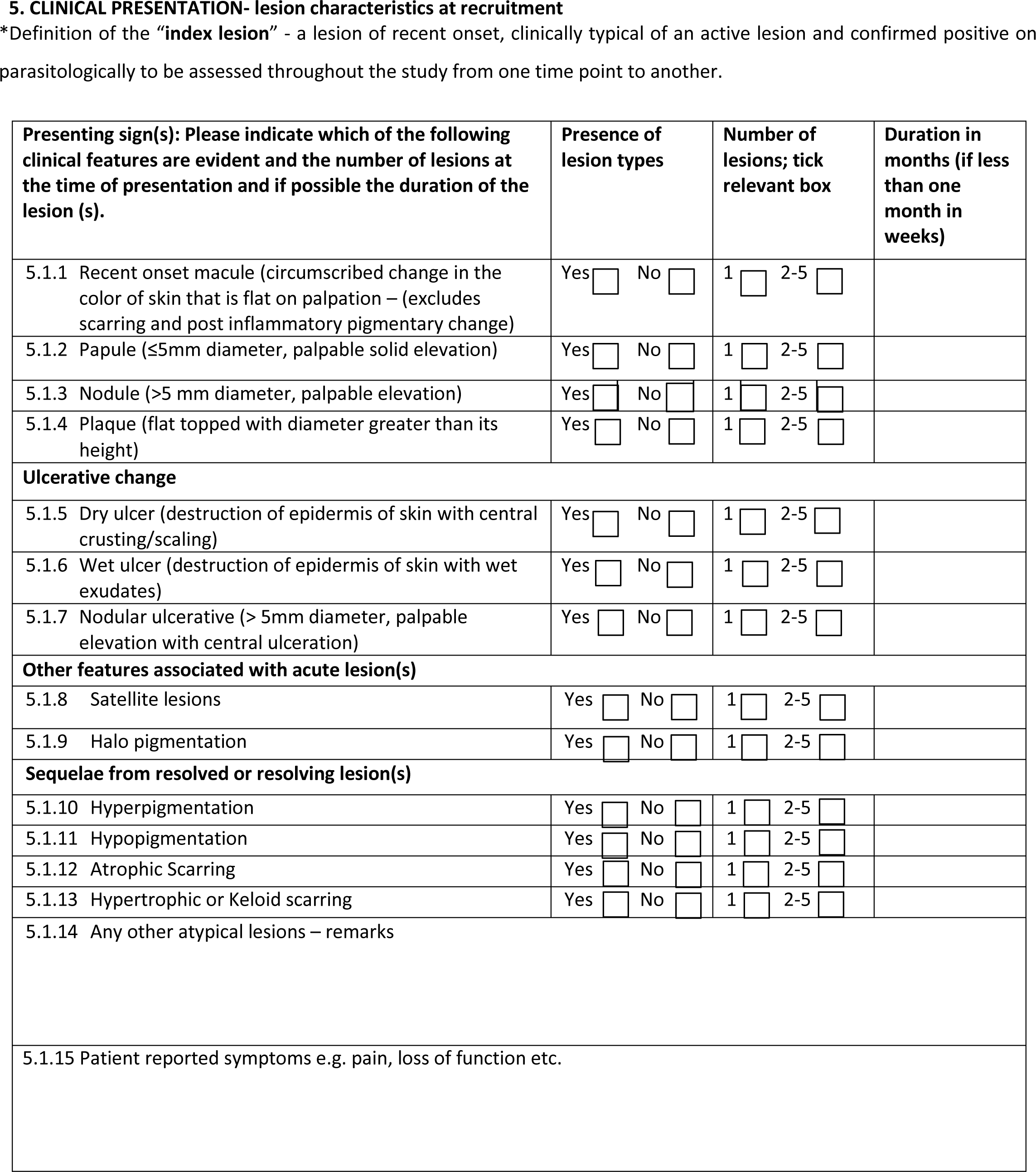

#### LOCALISED CUTANEOUS LEISHMANIASIS: BASELINE VISIT PRE-TREATMENT

6.1 Baseline biopsy and index lesion(s) for assessment

a. Please indicate on the figure below the site(s) of any biopsy(ies) taken and mark with a letter e.g. A
b. Draw the lesion(s) biopsied in the box and indicate where the biopsy has been taken (M= medial L = lateral)
c. Indicate features of the lesion(s) biopsied in the table below
d. Distinguish any index lesion(s) for assessment throughout the study from the biopsy sites on the figures, and mark with a different letter.

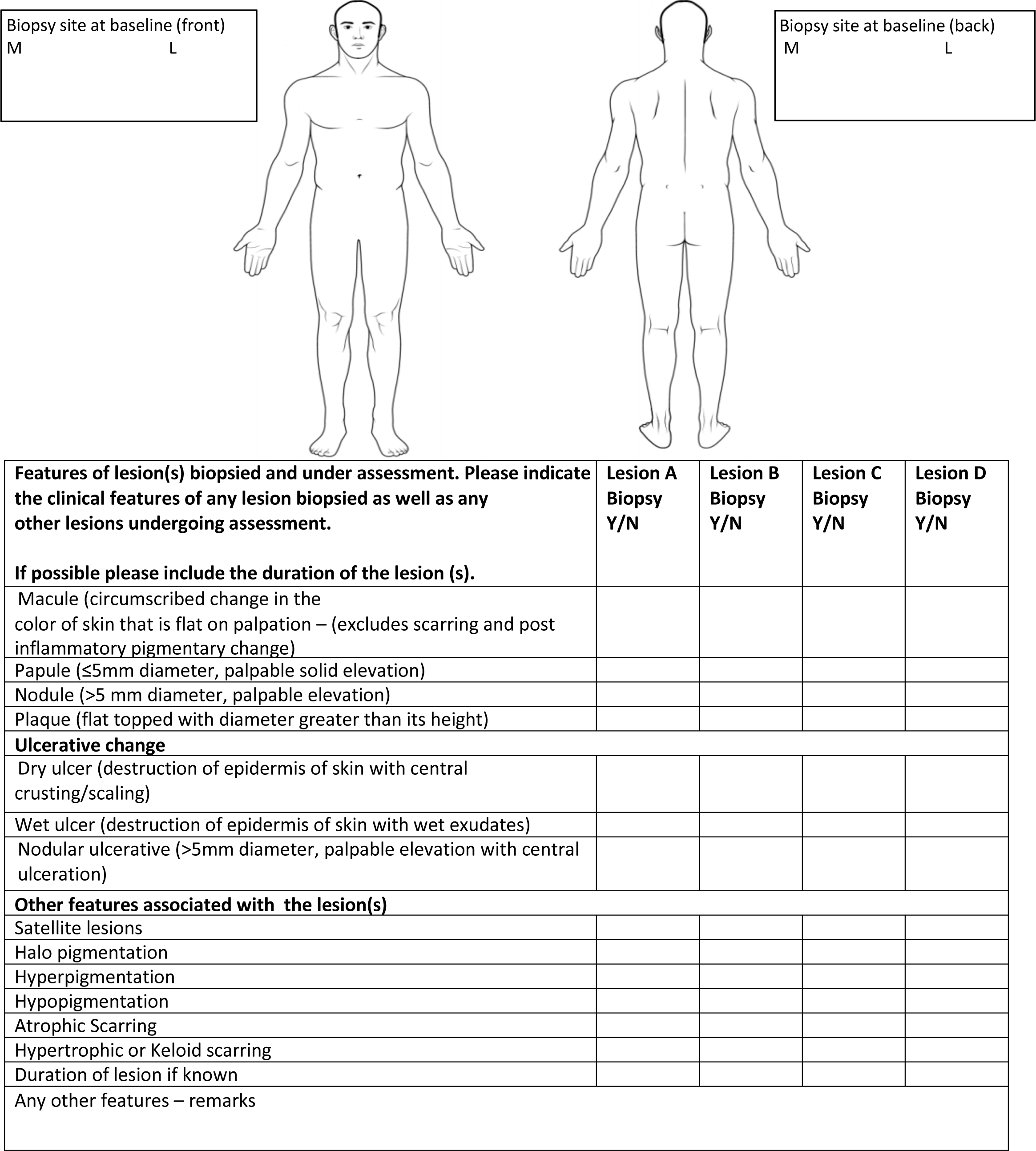

#### 6.2 OBJECTIVE ASSESSMENTS: BASELINE

**Area of Ulcer and Induration; See page 2 for instructions.**

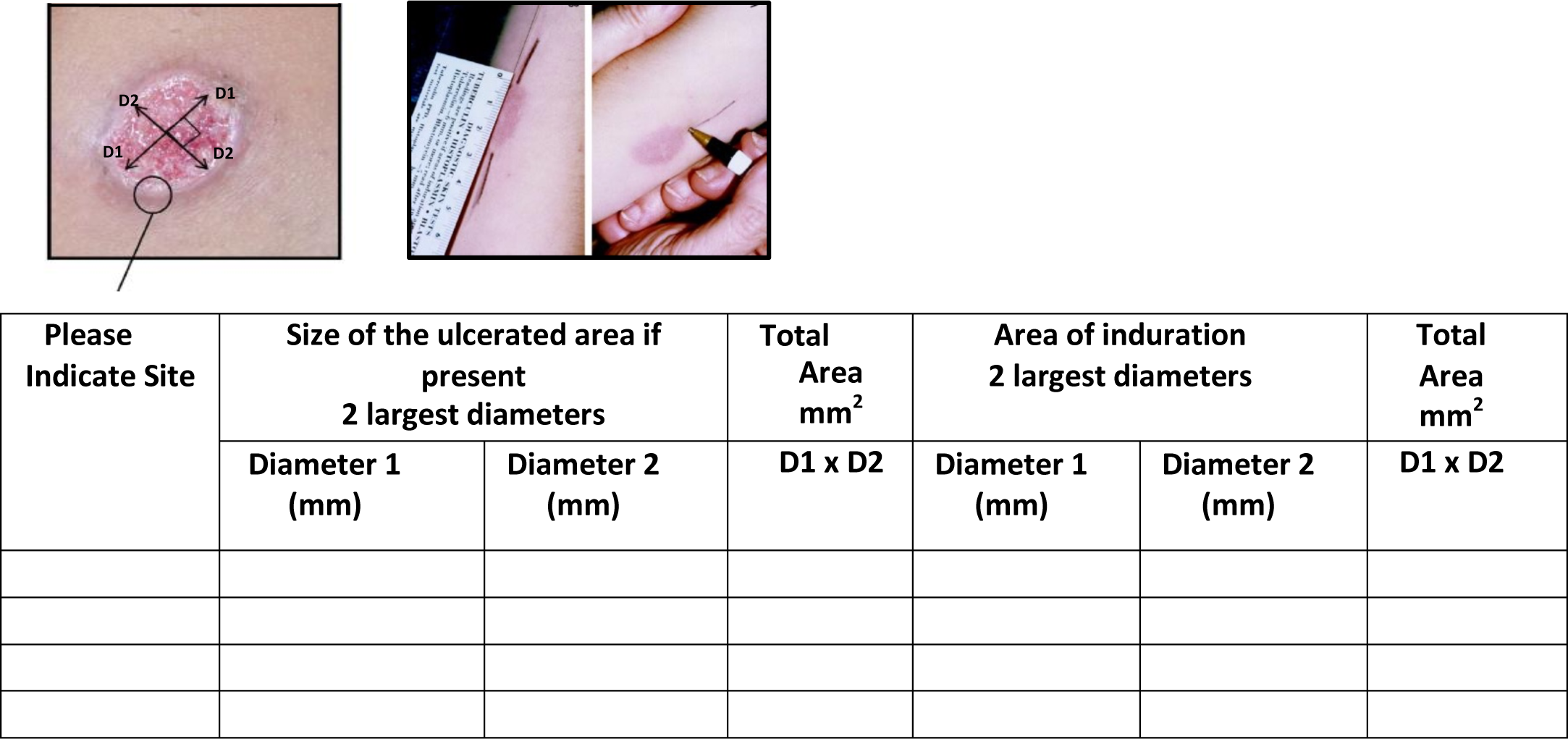

#### 6.3 SUBJECTIVE ASSESSMENTS: BASELINE

**Assessment using a palpability score**

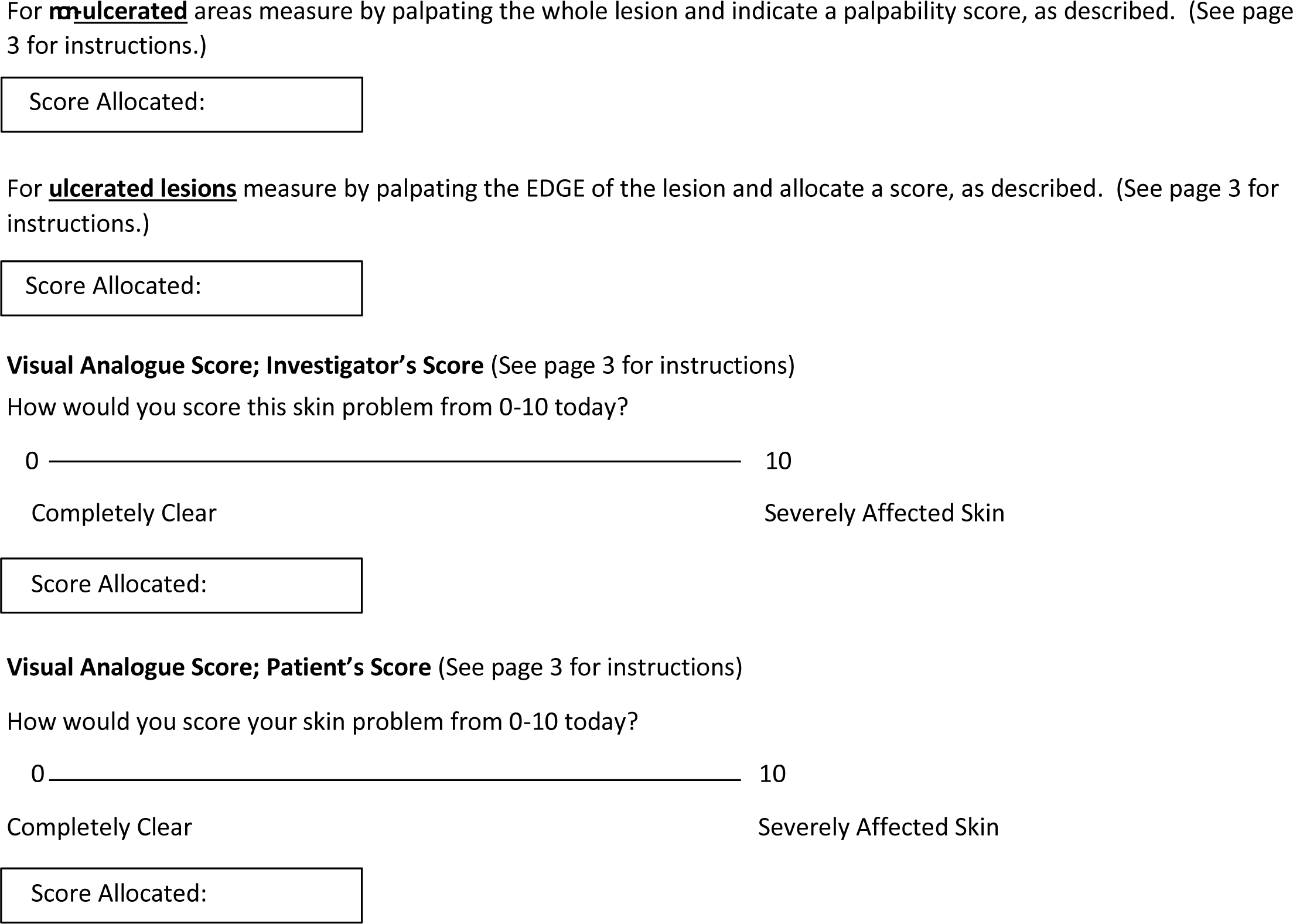

#### 6.4 TREATMENT EFFECT SCORES: BASELINE

**Investigator’s assessment of active disease post treatment**

**<u>Not required for Baseline visit</u>**

#### 6.5 SEQUELAE ASSESSMENTS: BASELINE

**Investigator Global Assessment of i & ii) Pigment change iii) Atrophic scars iv) Hypertrophic/ Keloid scars**

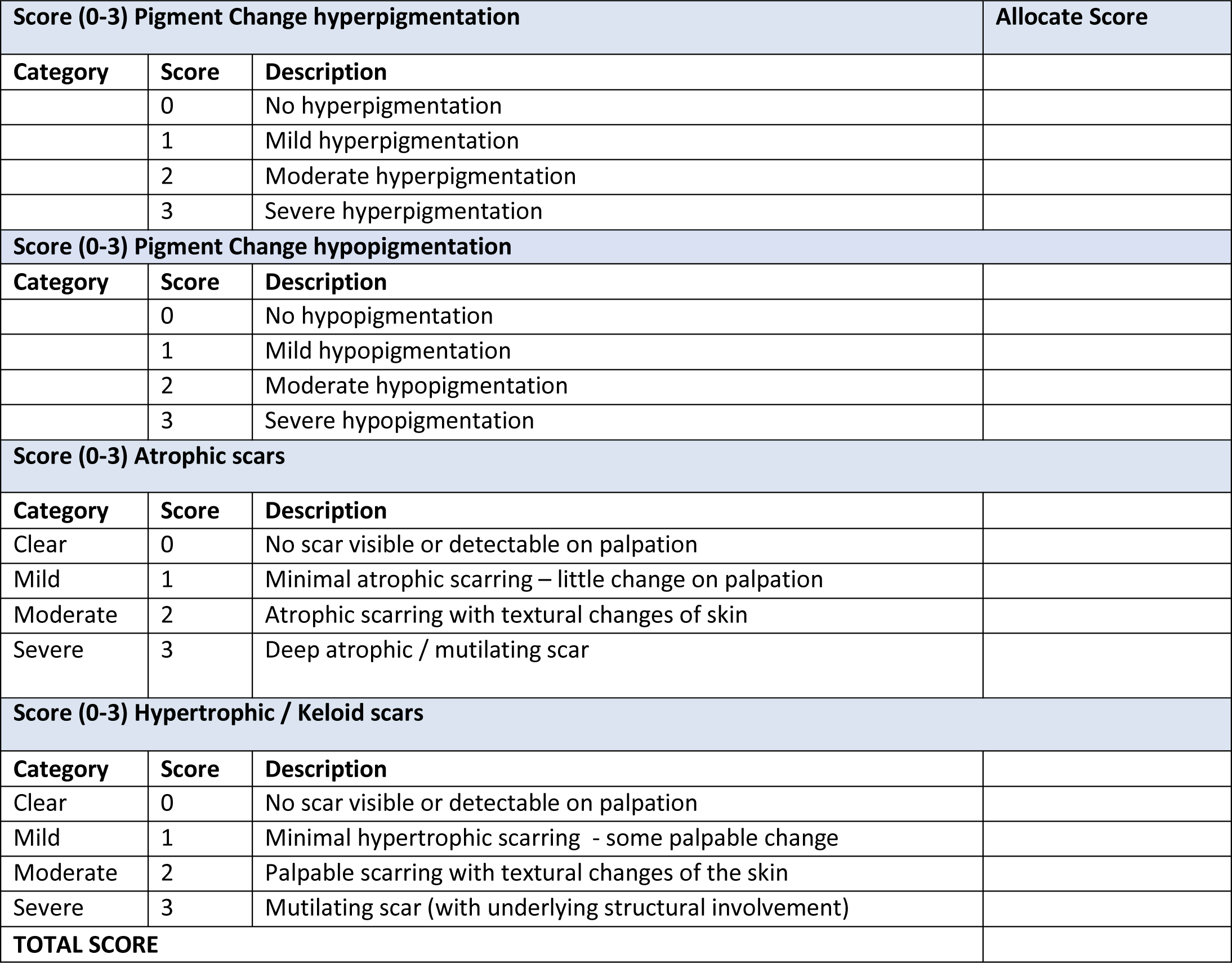

#### 6.6 HRQoL: BASELINE

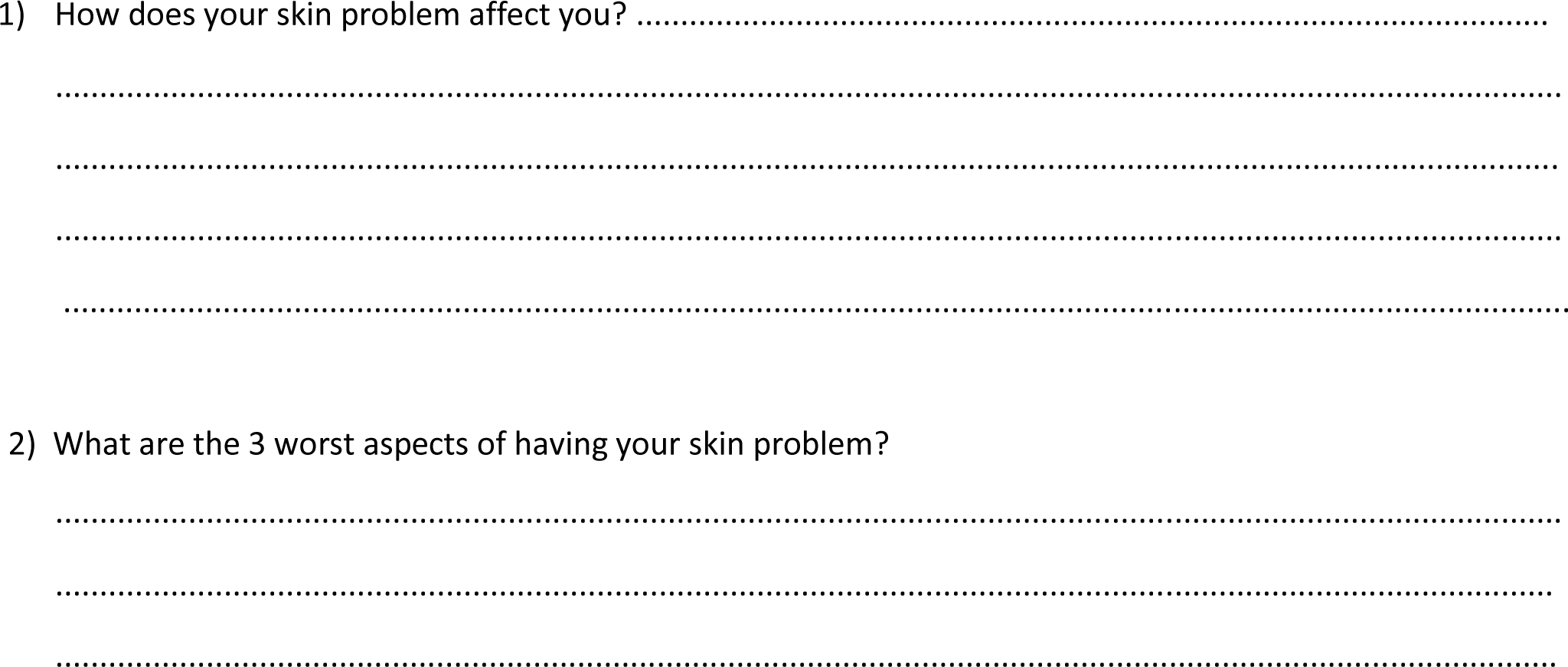

#### 6.7 SUMMARY OF SCORES: BASELINE

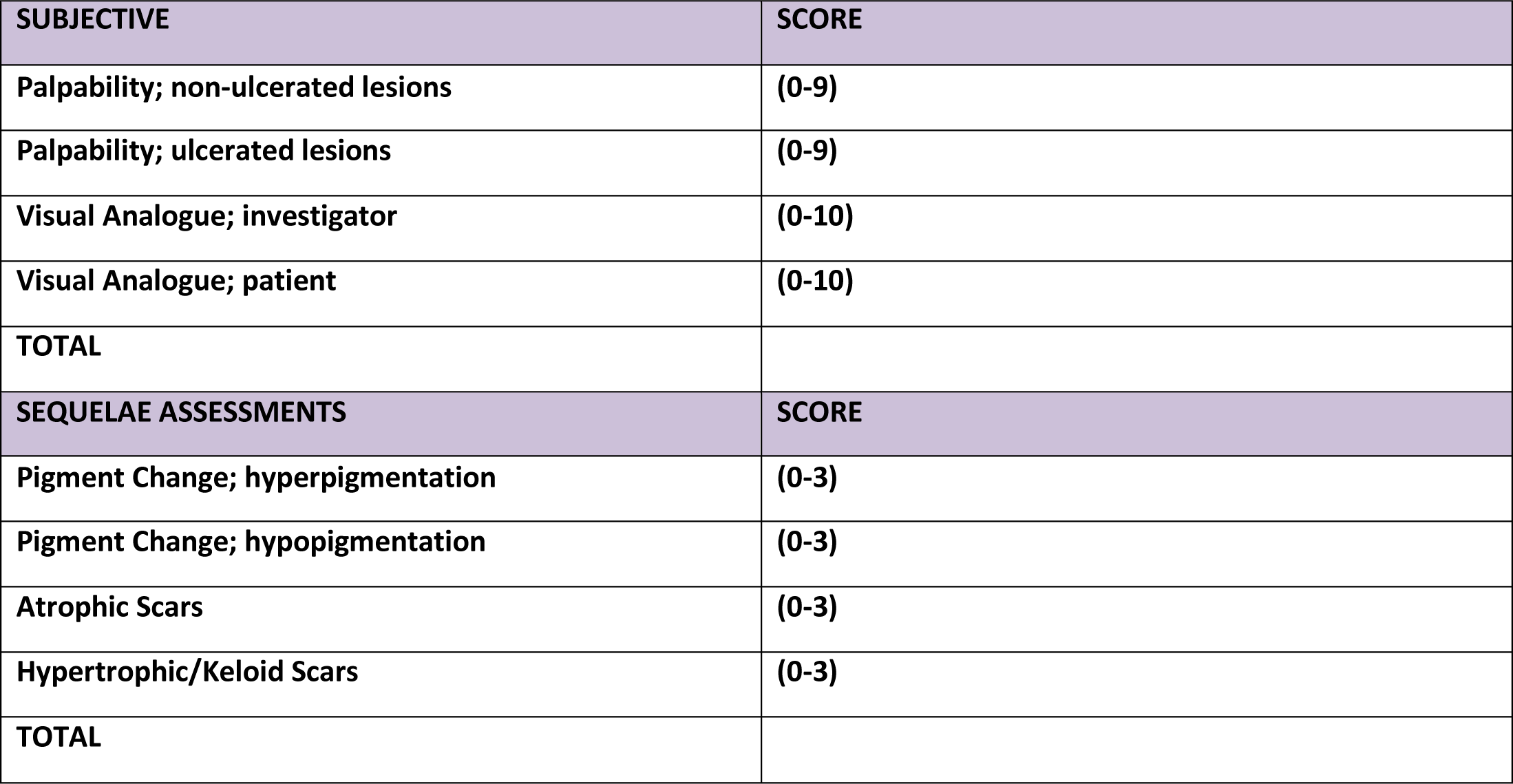

### 7. LOCALISED CUTANEOUS LEISHMANIASIS: 4 WEEKS

#### 7.1. FOUR WEEK BIOPSY AND INDEX LESION(S) FOR ASSESSMENT

a. Please indicate on the figure below the site(s) of any biopsy(ies) taken and mark with a letter e.g. A
b. Draw the lesion(s) biopsied in the box and indicate where the biopsy has been taken (M= medial L = lateral)
c. Indicate features of the lesion(s) biopsied in the table below
d. Distinguish any index lesion(s) for assessment throughout the study from the biopsy sites on the figures, and mark with a different letter.

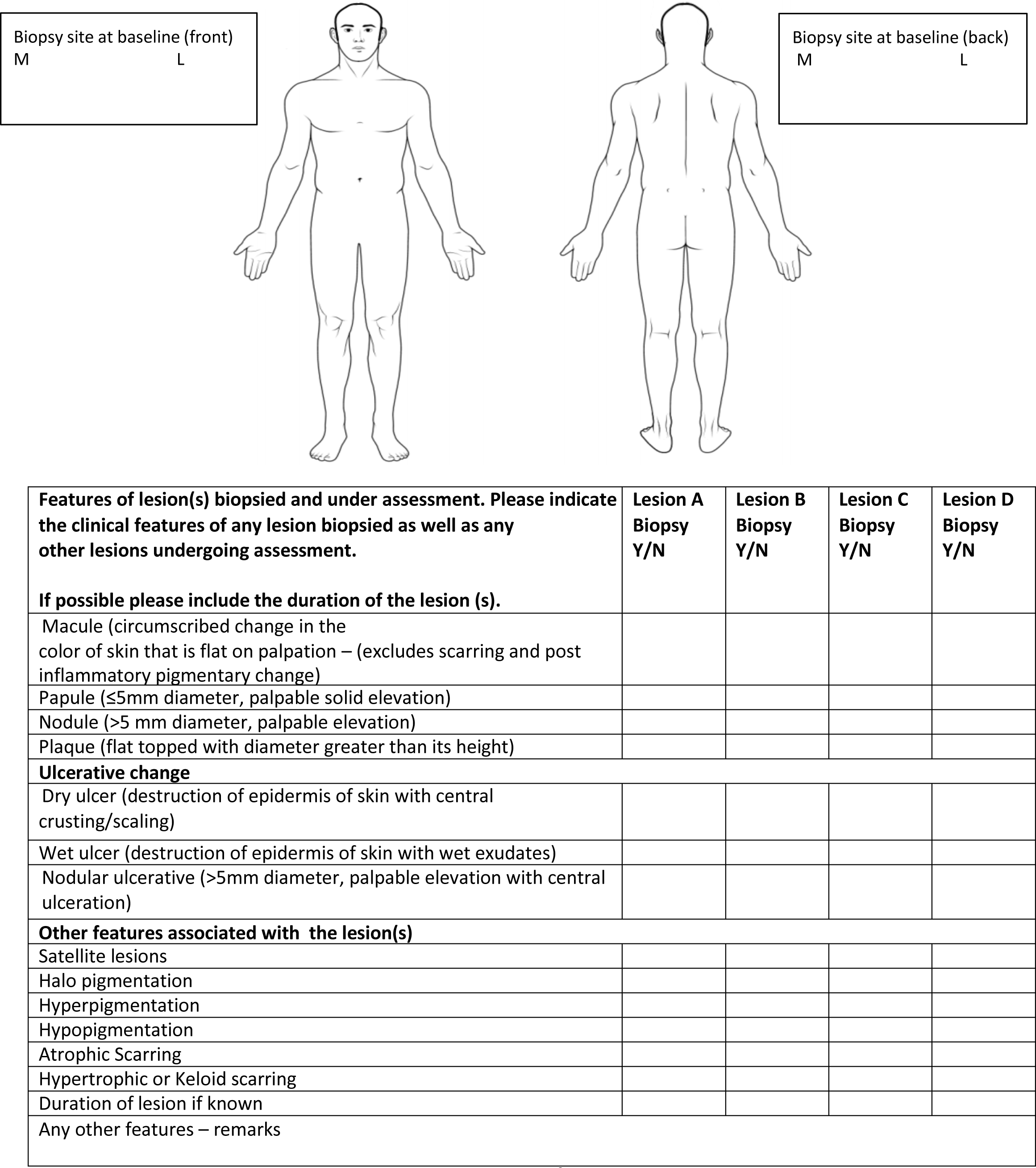

#### 7.2. OBJECTIVE ASSESSMENTS OF INDEX LESION(S): 4 WEEKS

**Area of Ulcer and Induration. (See page 2 for instructions)**

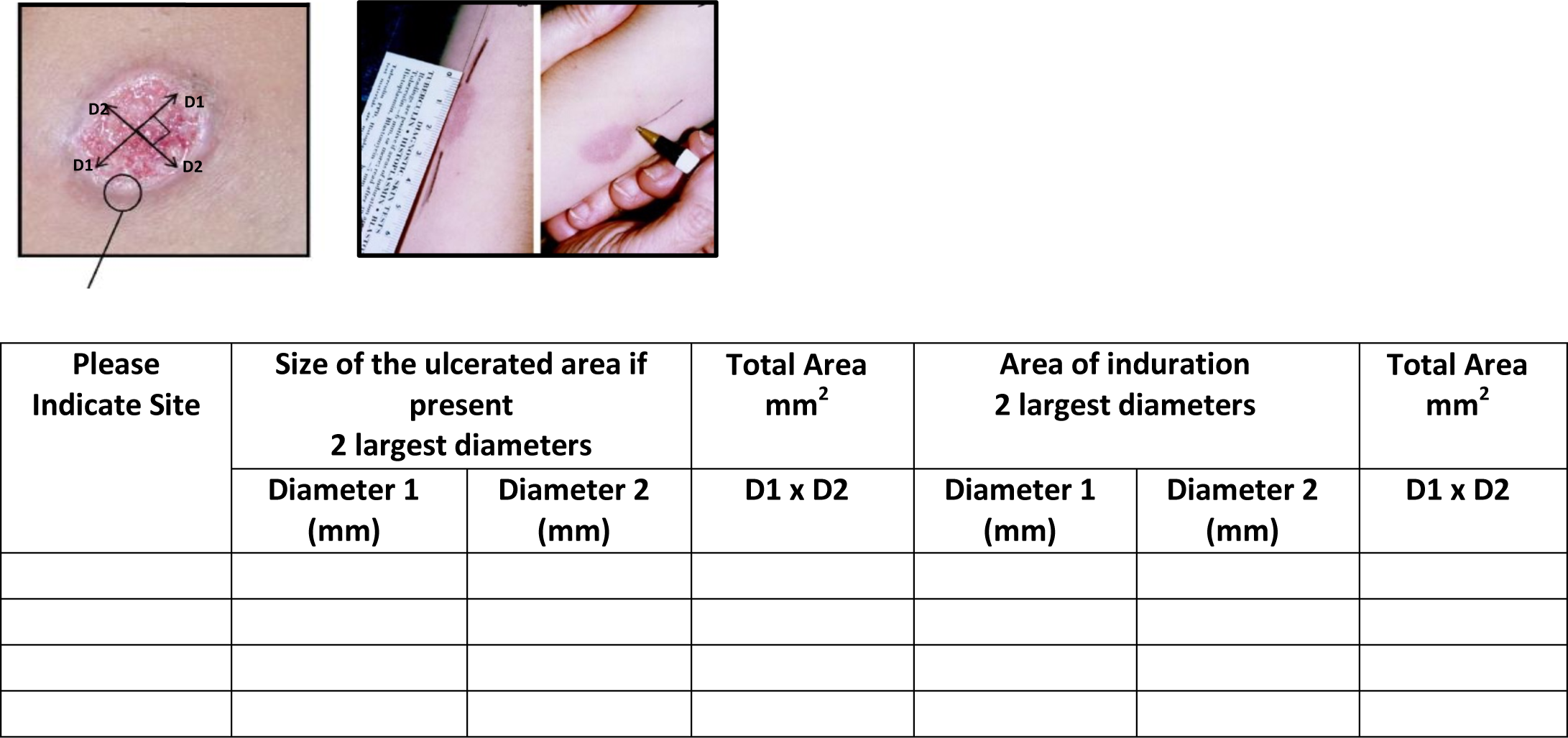

#### 7.3. SUBJECTIVE ASSESSMENTS: 4 WEEKS

**Assessment using a palpability score**

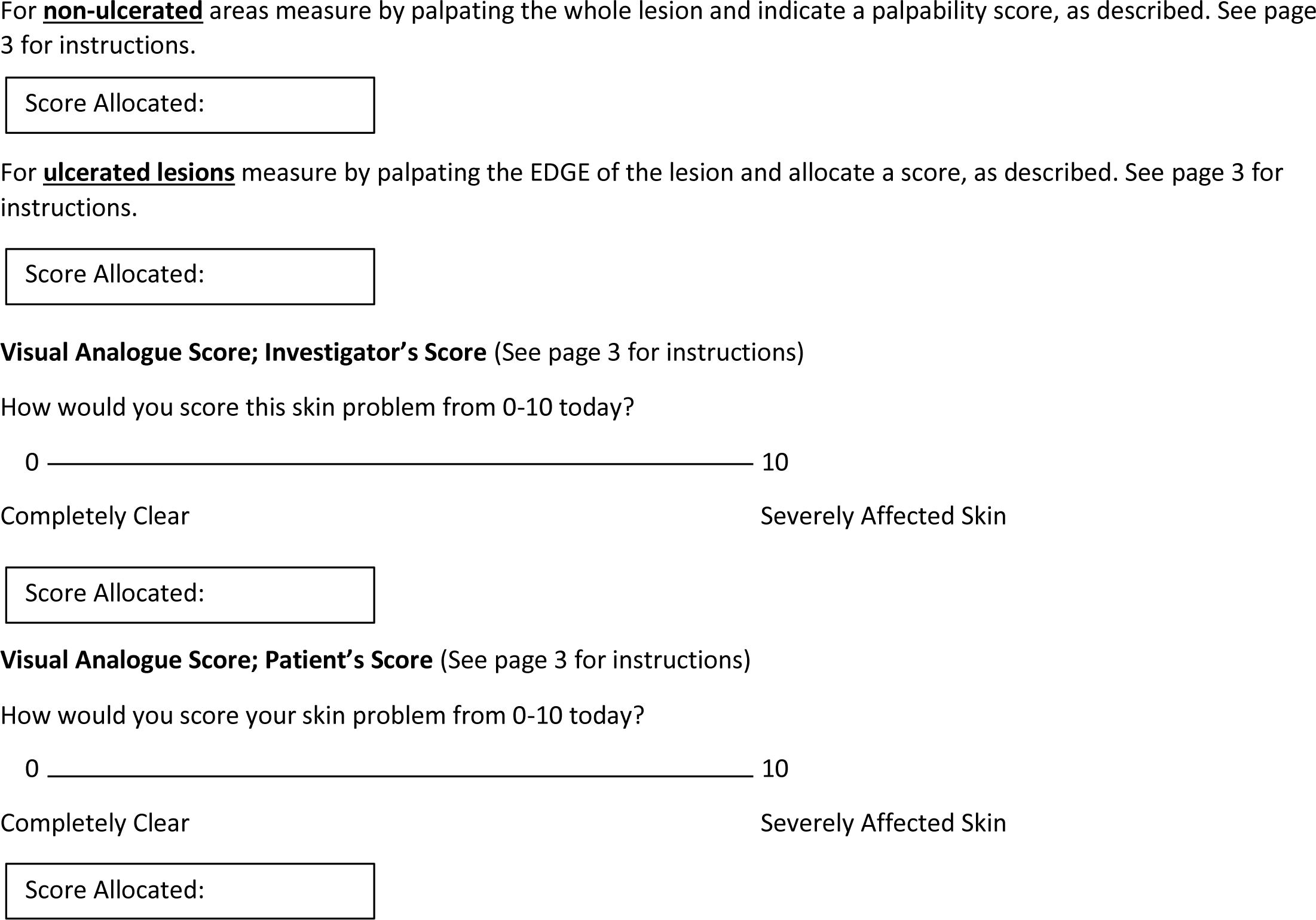

#### 7.4. TREATMENT EFFECT SCORES: 4 WEEKS

**Investigator Global assessment of active disease post treatment**

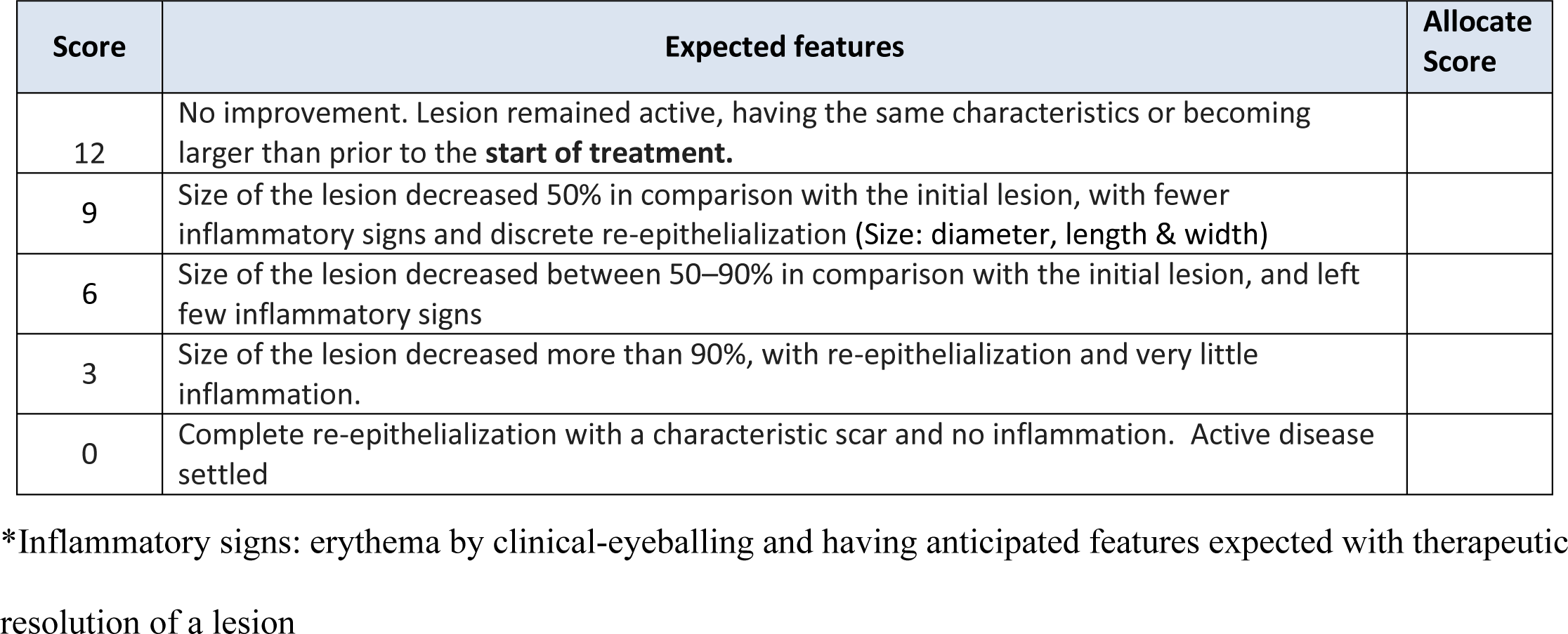

#### 7.5. SEQUELAE ASSESSMENTS: 4 WEEKS

**Investigator Global Assessment of i & ii) Pigment change iii) Atrophic scars iv) Hypertrophic/ Keloid scars**

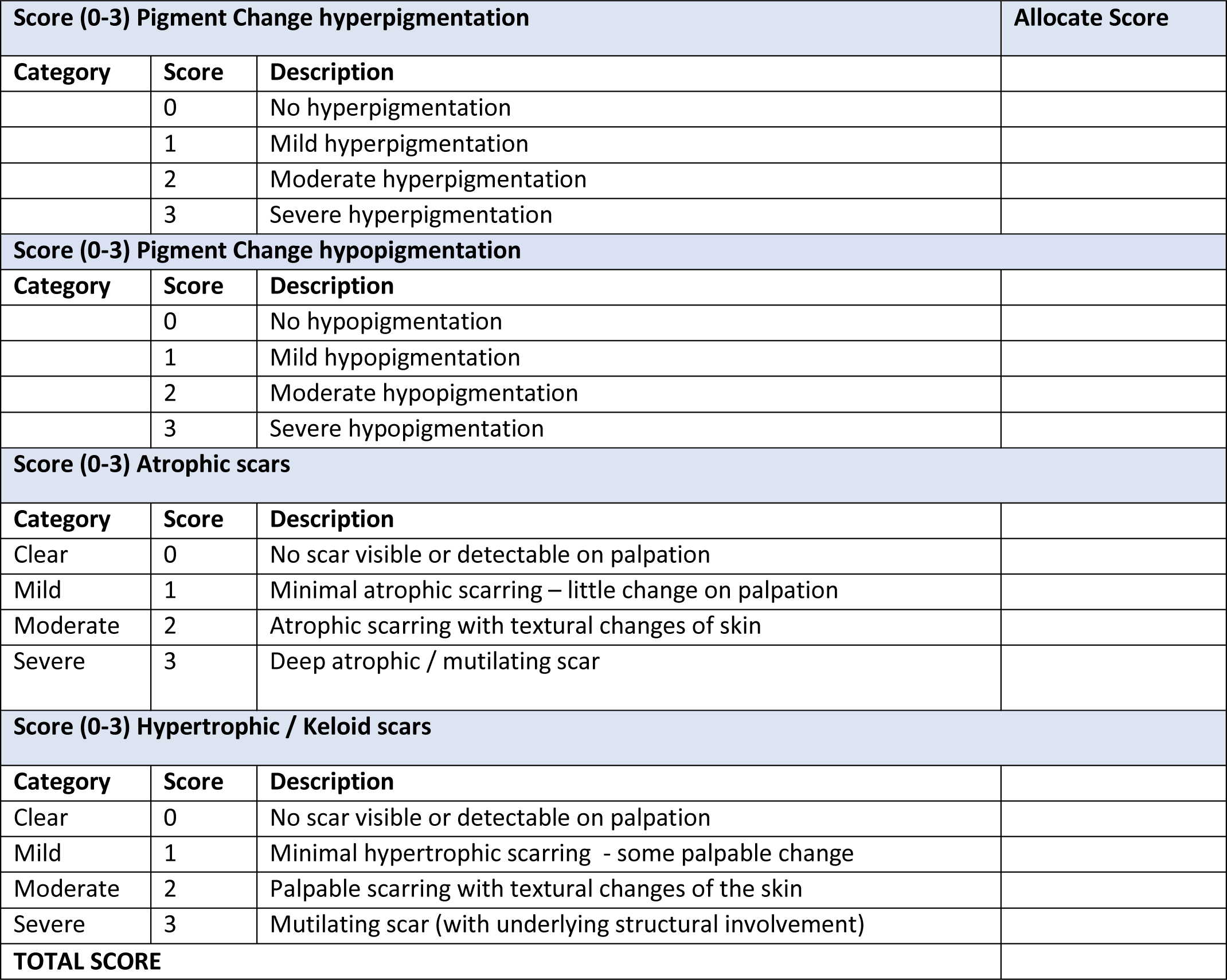

#### 7.6. HRQoL: 4 WEEKS

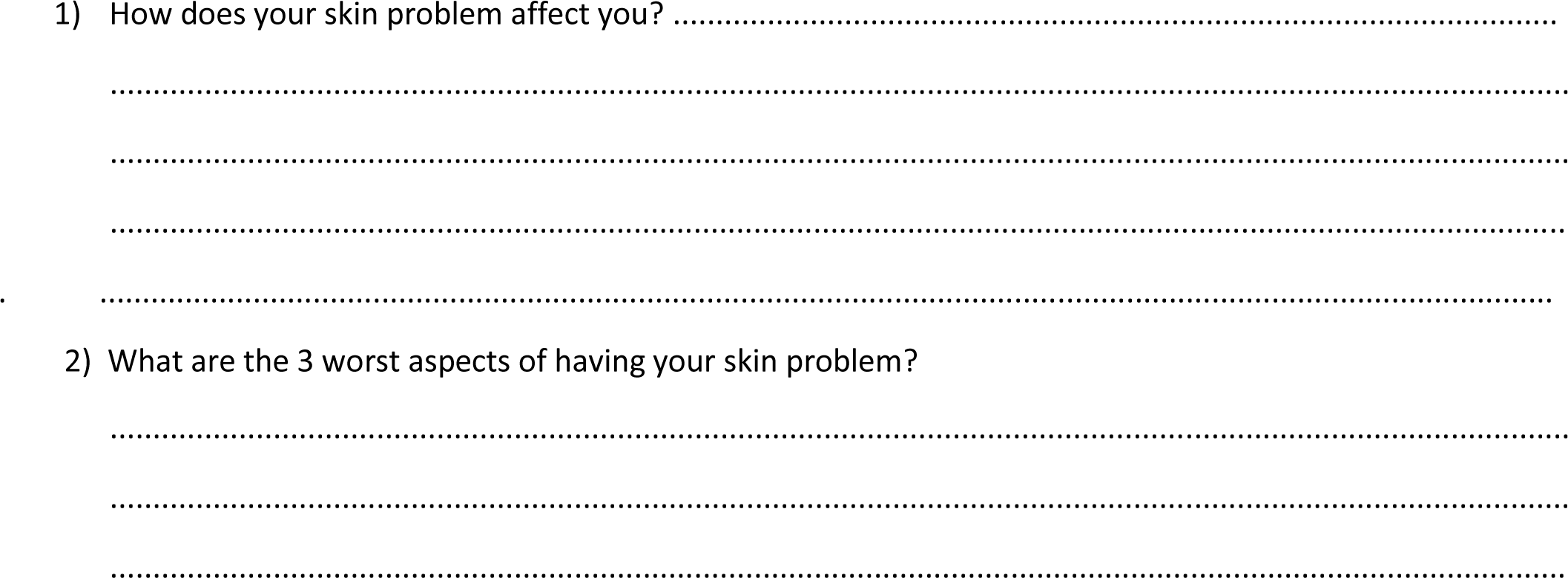

#### 7.7. SUMMARY OF SCORES: 4 WEEKS

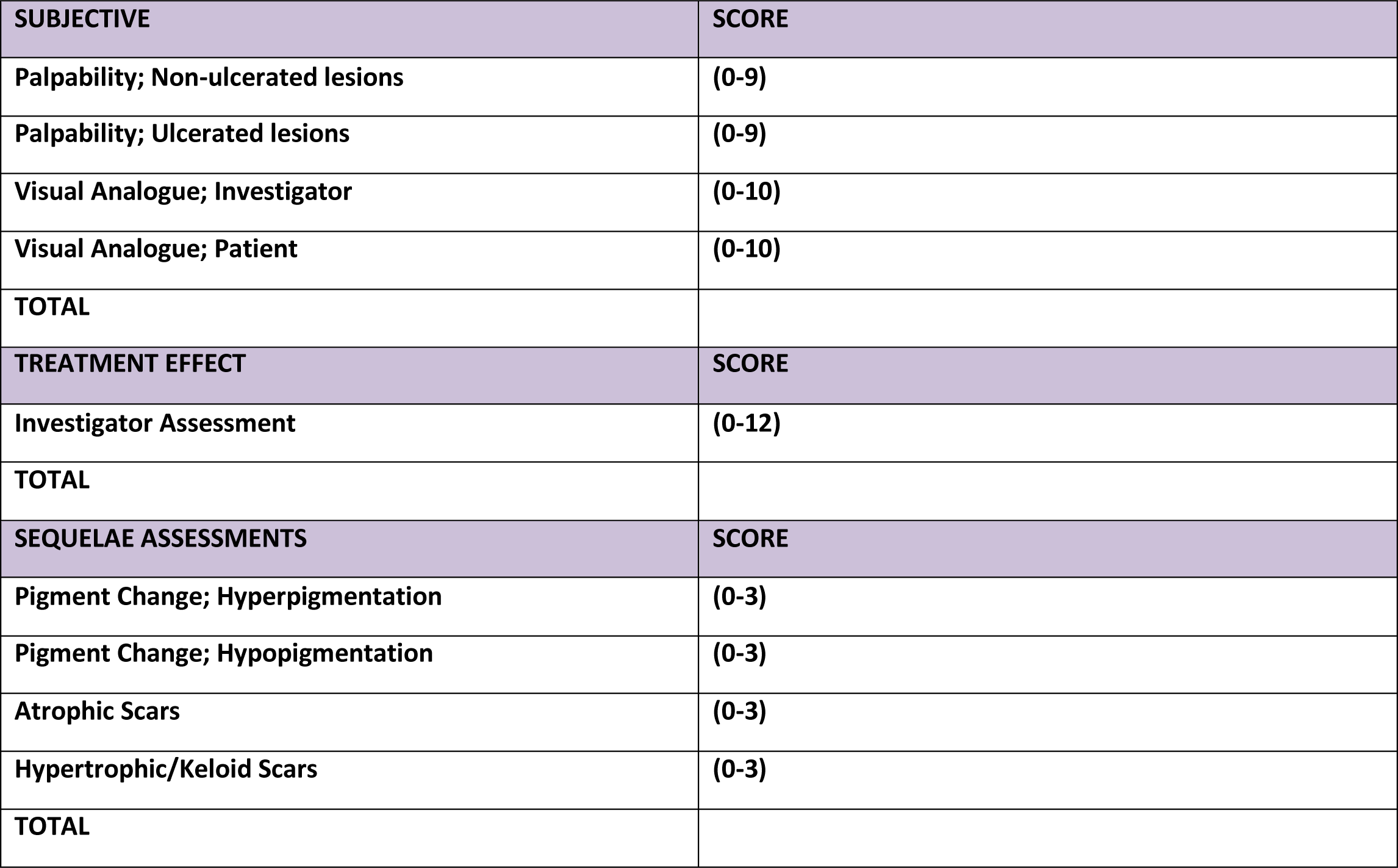

### 8. LOCALISED CUTANEOUS LEISHMANIASIS: 3 MONTHS

#### 8.1. 3 MONTH BIOPSY (IF TAKEN) AND INDEX LESION(S) FOR ASSESSMENT

a. Please indicate on the figure below the site(s) of any biopsy(ies) taken and mark with a letter e.g. A
b. Draw the lesion(s) biopsied in the box and indicate where the biopsy has been taken (M= medial L = lateral)
c. Indicate features of the lesion(s) biopsied in the table below
d. Distinguish any index lesion(s) for assessment throughout the study from the biopsy sites on the figures, and mark with a different letter.

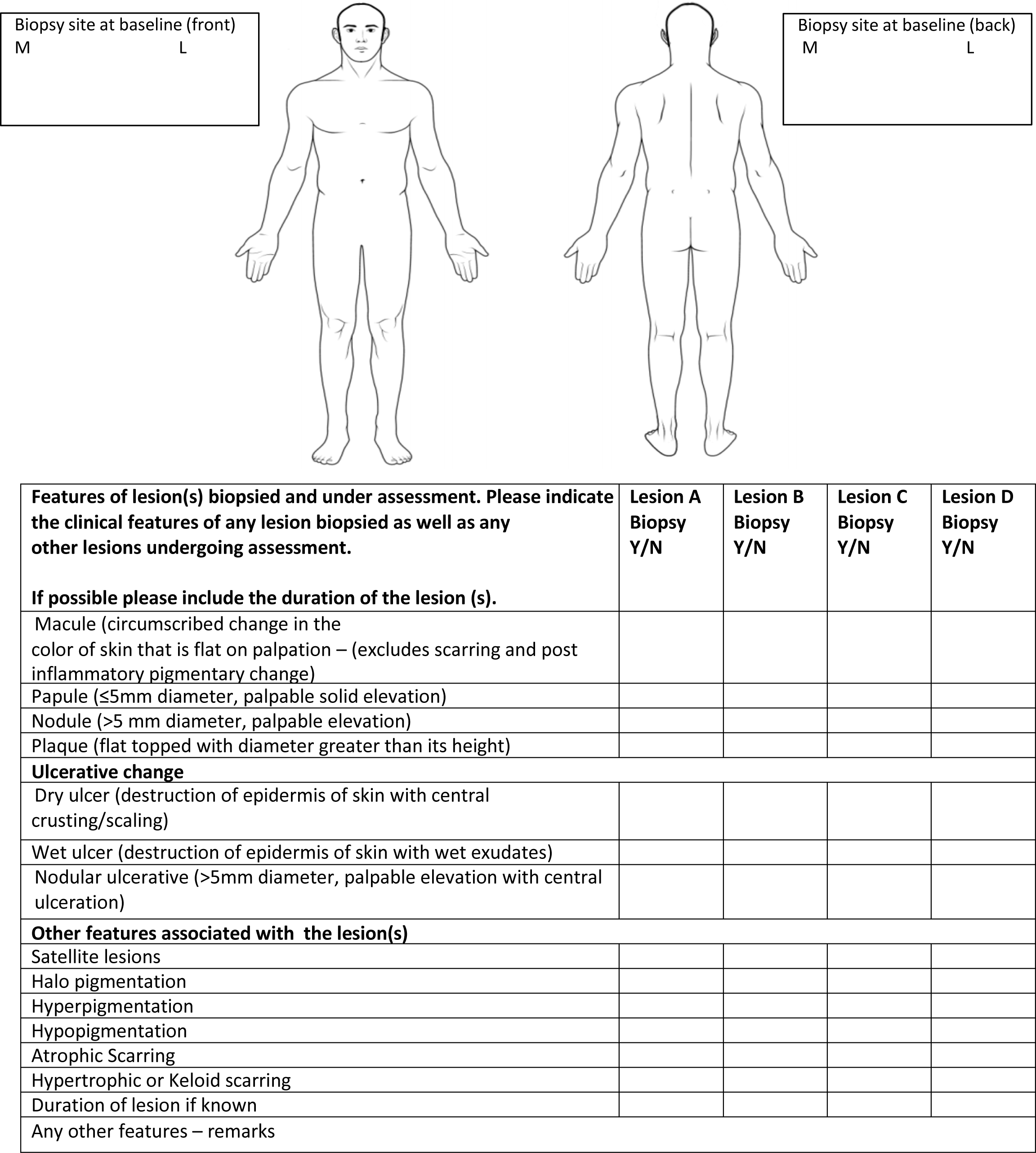

#### 8.2. OBJECTIVE ASSESSMENTS: 3 MONTHS

**Area of Ulcer and Induration (See page 2 for instructions)**

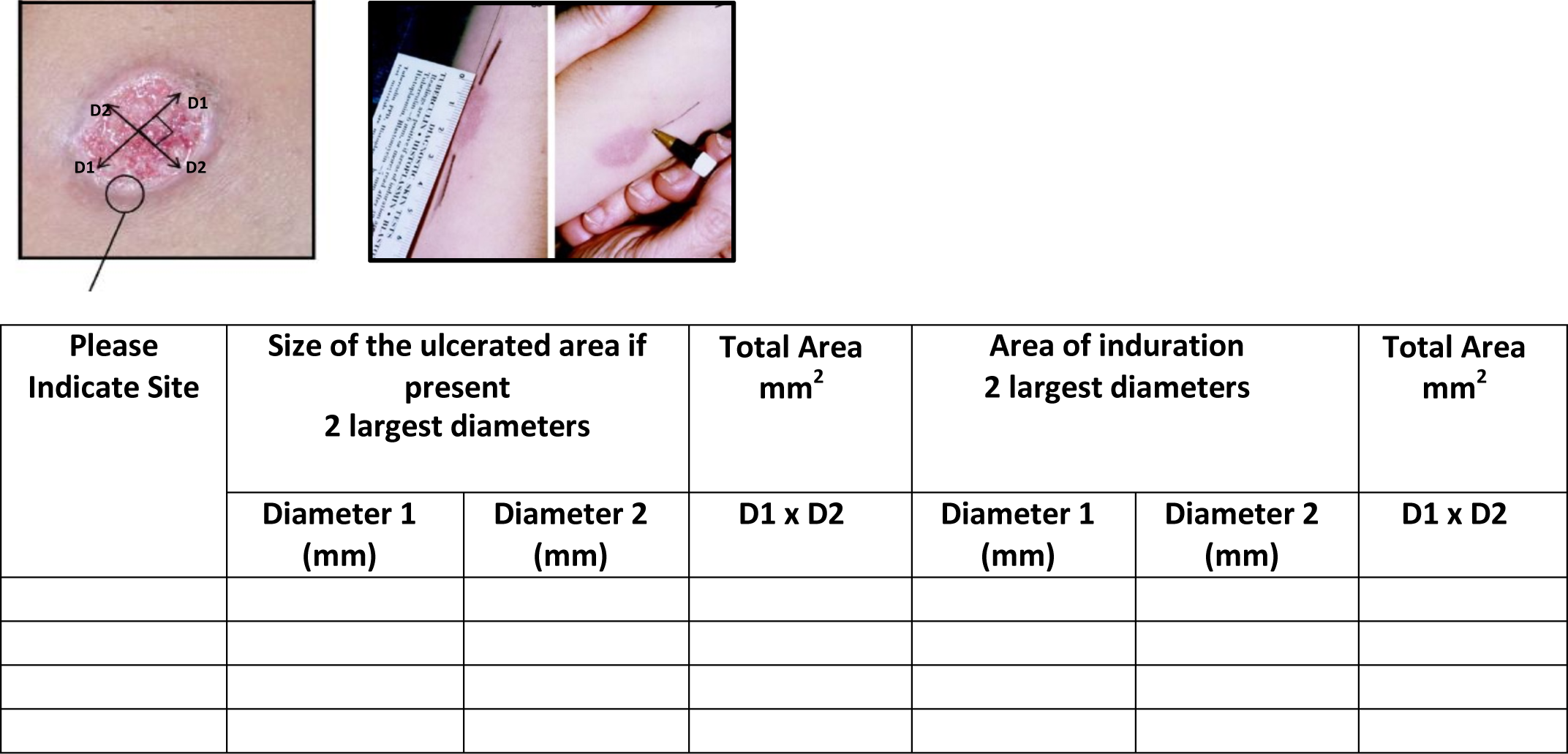

#### 8.3. SUBJECTIVE ASSESSMENTS: 3 MONTHS

**Assessment using a palpability score**

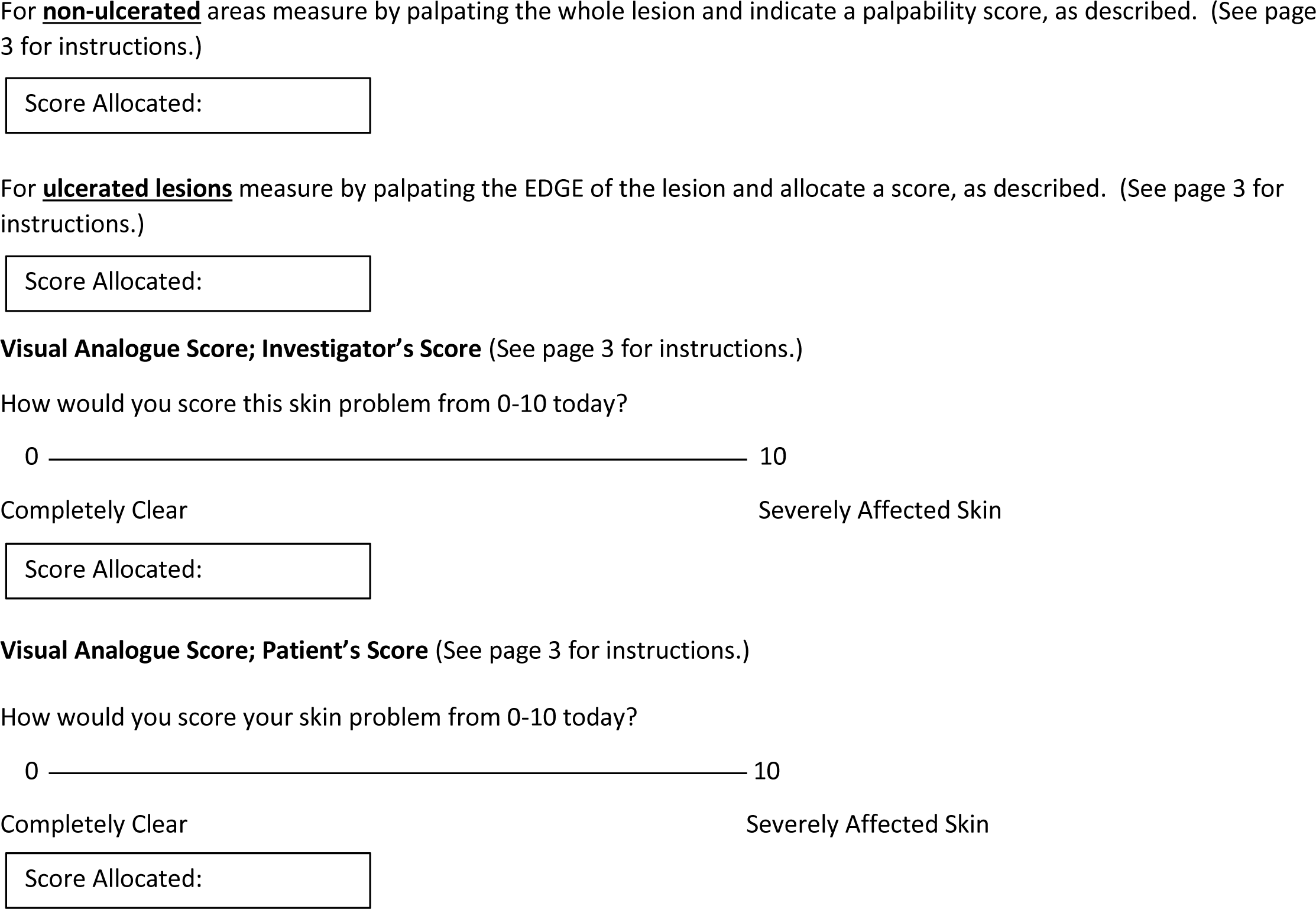

#### 8.4. TREATMENT EFFECT SCORES: 3 MONTHS

**Investigator Global assessment of active disease post treatment**

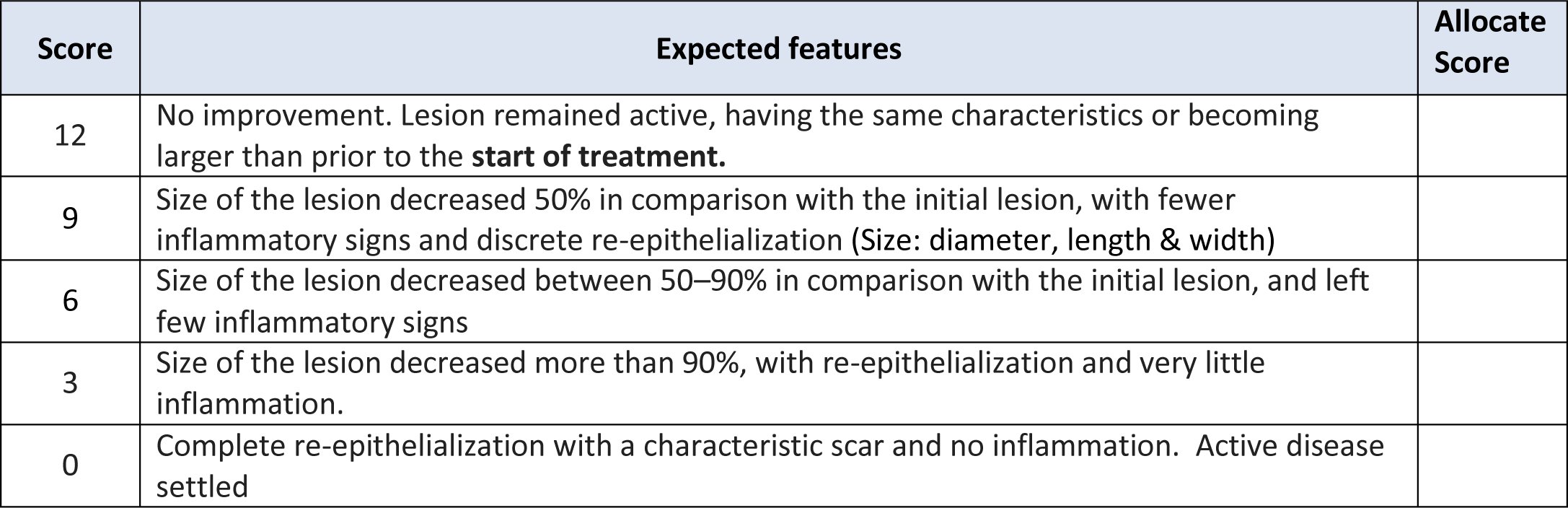

#### 8.5. SEQUELAE ASSESSMENTS: 3 MONTHS

**Investigator Global Assessment of i & ii) Pigment change iii) Atrophic scars iv) Hypertrophic/ Keloid scars**

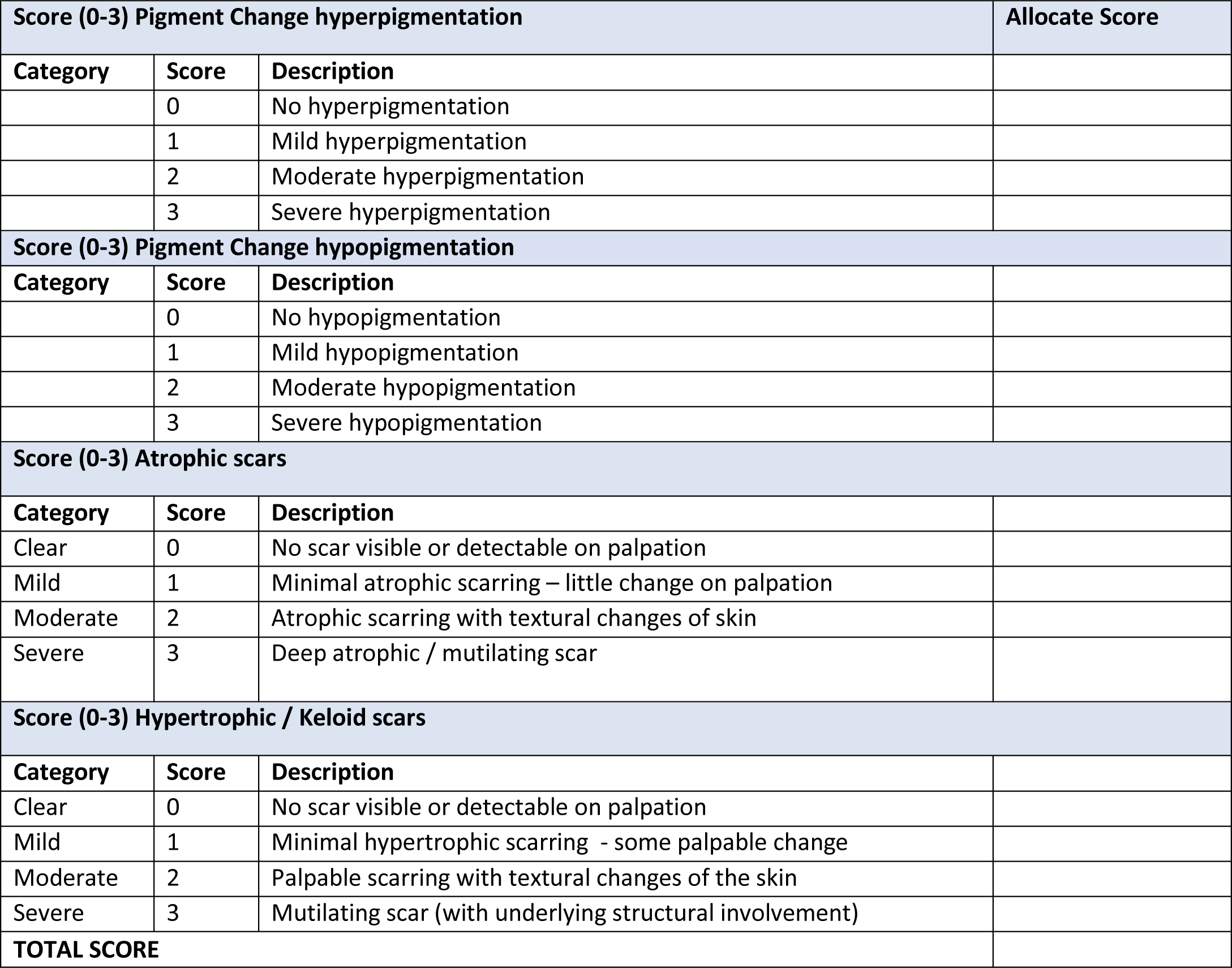

#### 8.6. MEASURING HRQoL: 3 MONTHS

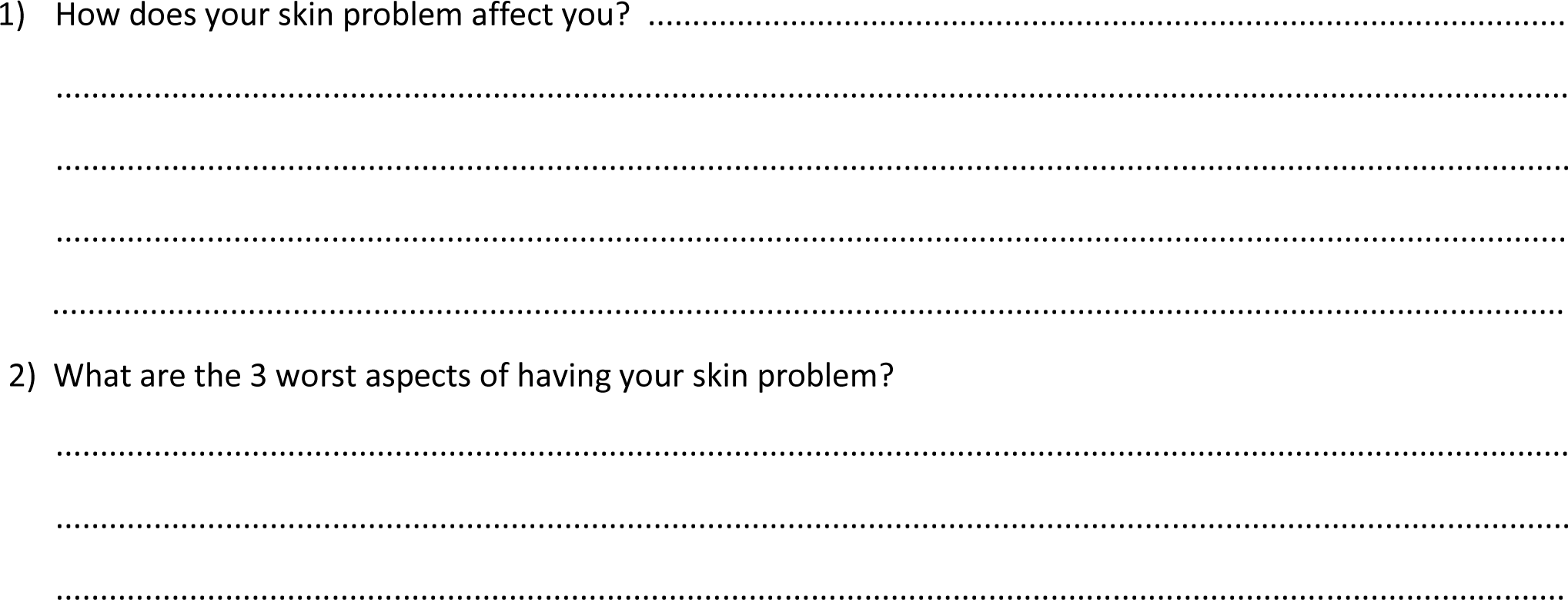

#### 8.7. SUMMARY OF SCORES: 3 MONTHS

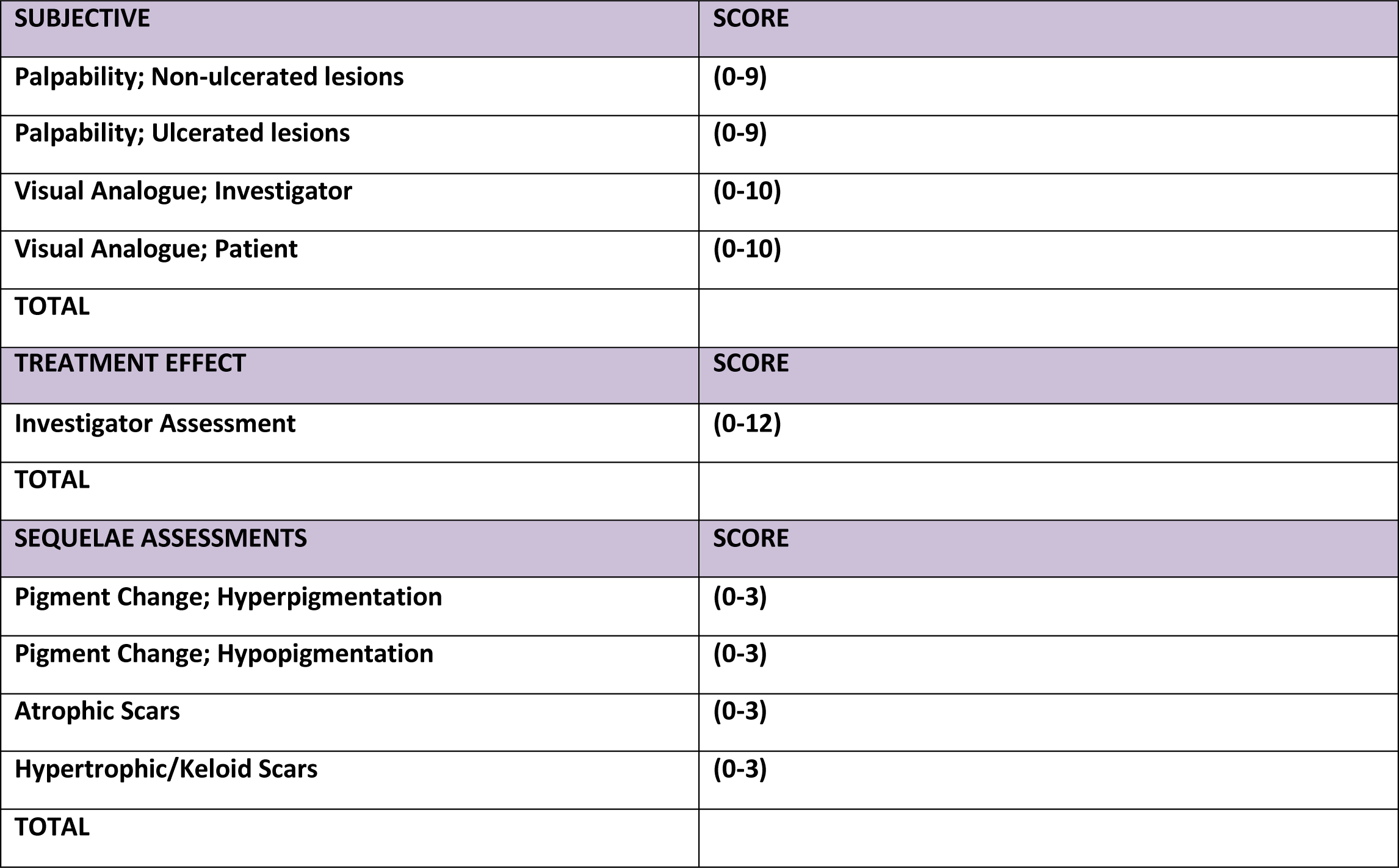

### 9. LOCALISED CUTANEOUS LEISHMANIASIS: 6 MONTHS

9.1 Baseline biopsy and index lesion(s) for assessment

a. Please indicate on the figure below the site(s) of any biopsy(ies) taken and mark with a letter e.g. A
b. Draw the lesion(s) biopsied in the box and indicate where the biopsy has been taken (M= medial L = lateral)
c. Indicate features of the lesion(s) biopsied in the table below
d. Distinguish any index lesion(s) for assessment throughout the study from the biopsy sites on the figures, and mark with a different letter.

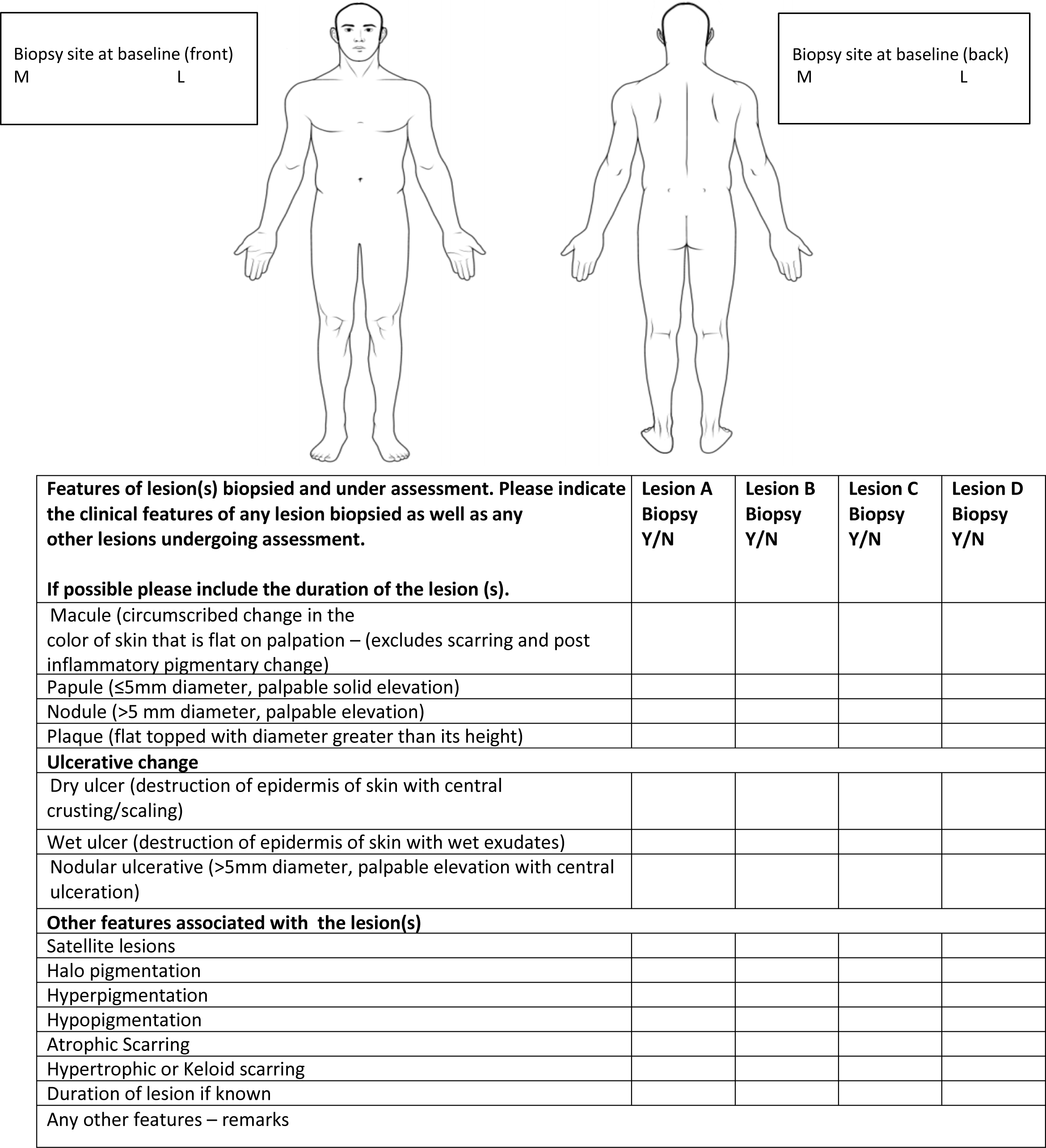

#### 9.2. OBJECTIVE ASSESSMENTS: 6 MONTHS

**Area of Ulcer and Induration; See page 2 for instructions**

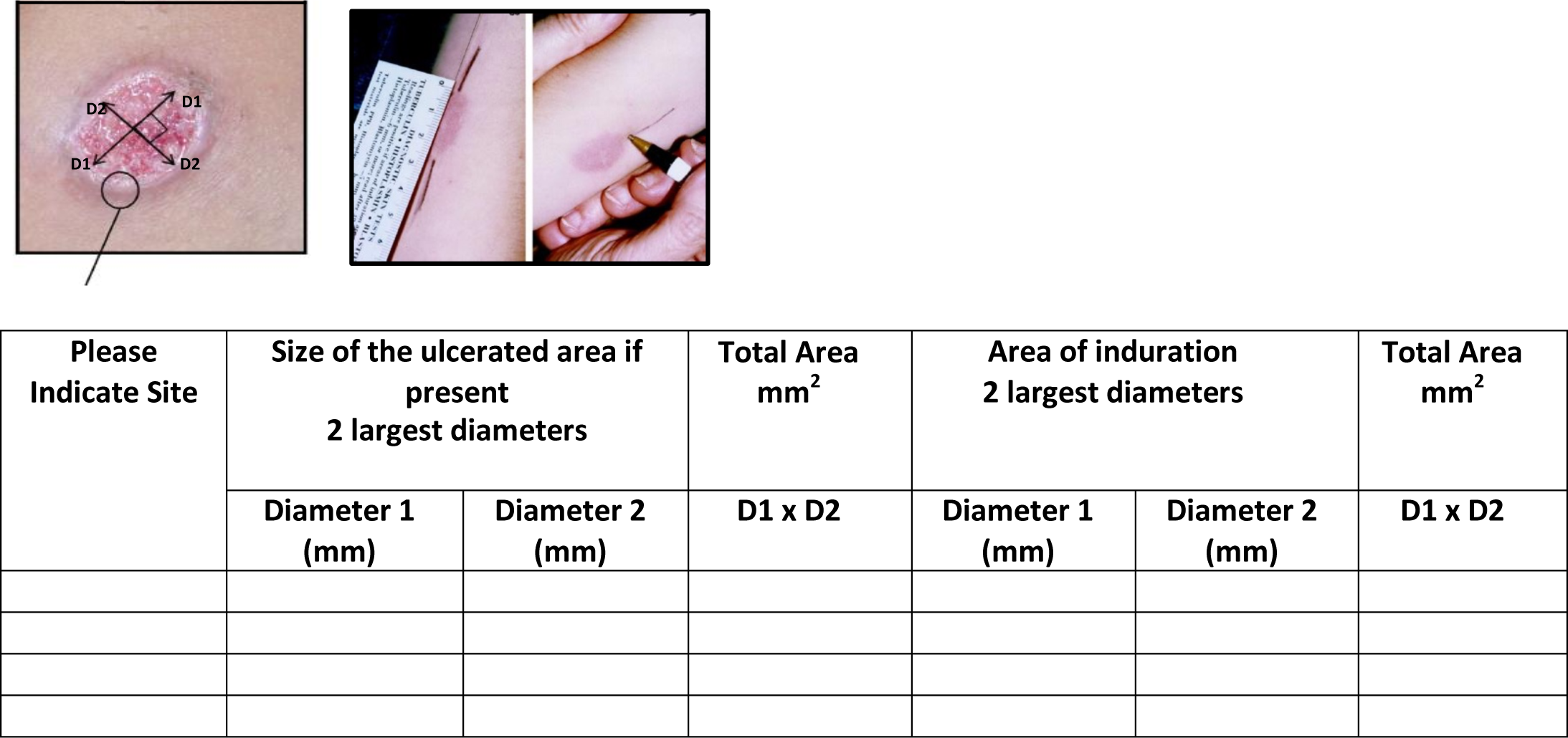

#### 9.3. SUBJECTIVE ASSESSMENTS: 6 MONTHS

**Assessment using a palpability score**

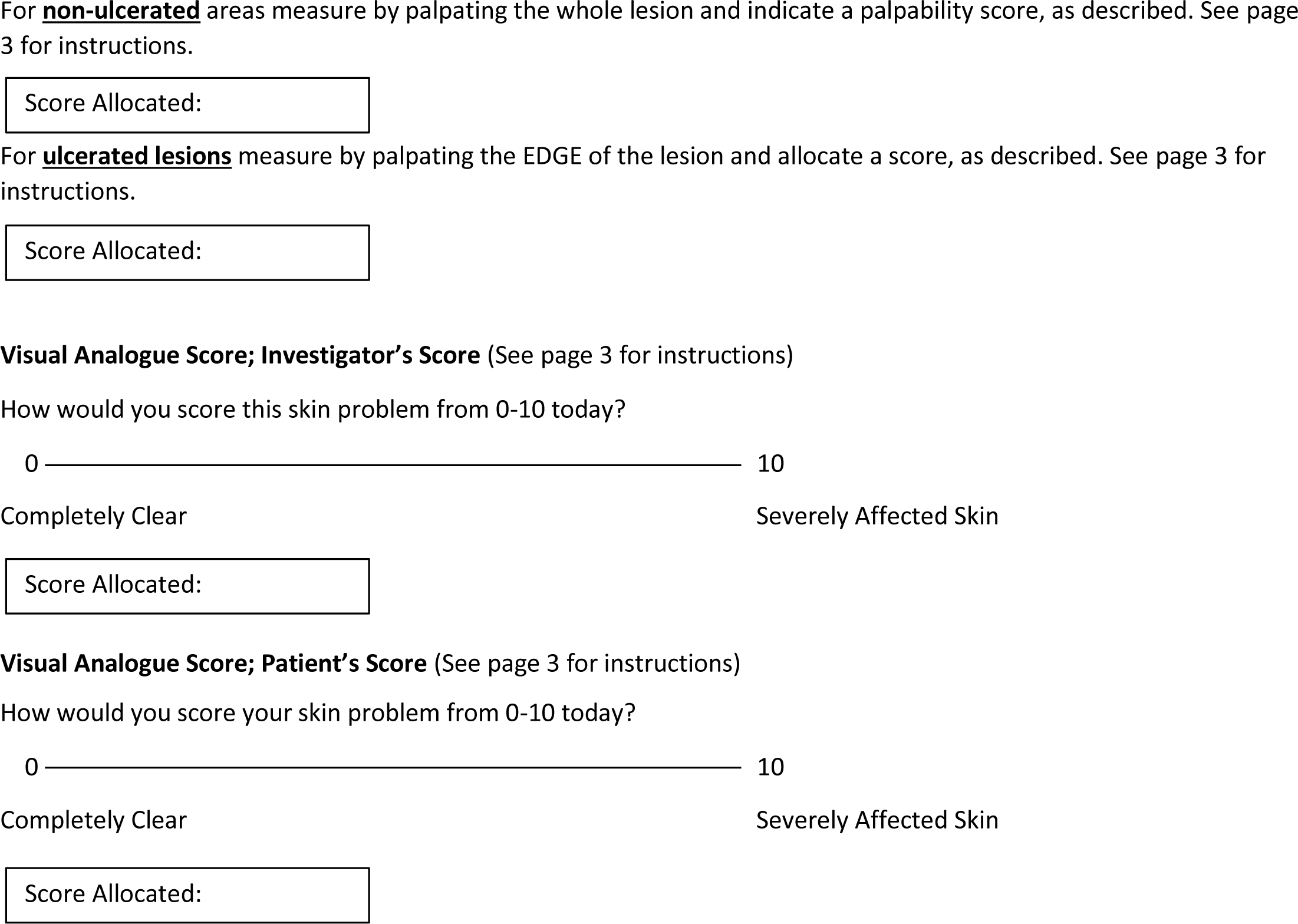

#### 9.4. TREATMENT EFFECT SCORES: 6 MONTHS

**Investigator Global assessment of active disease post treatment**

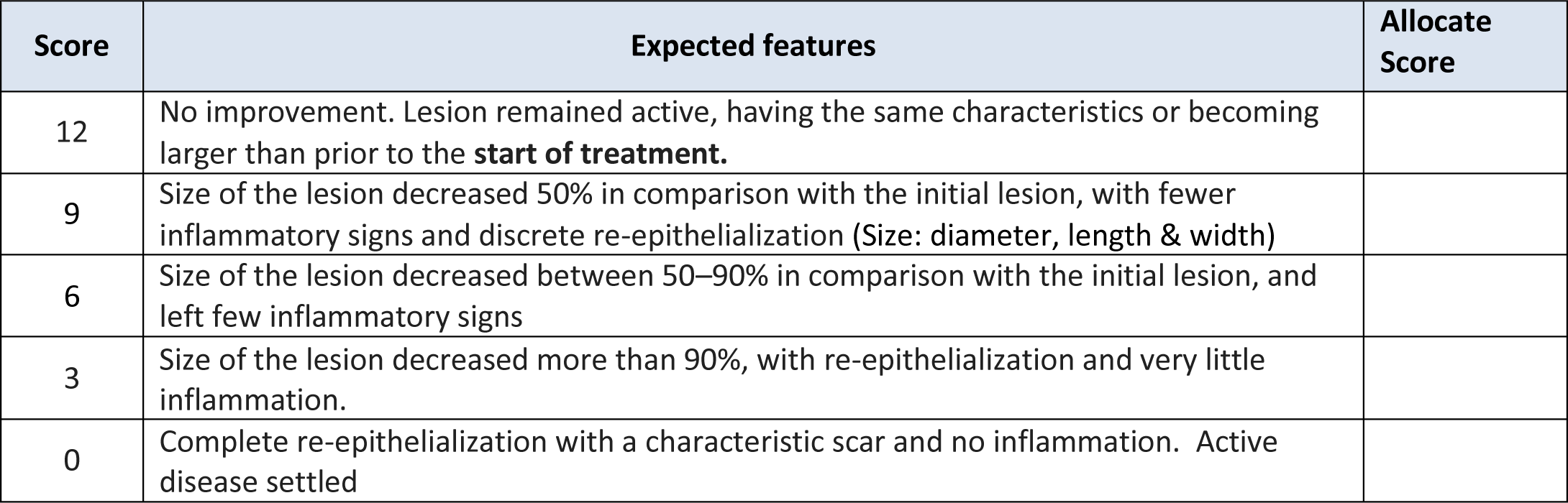

#### 9.5. SEQUELAE ASSESSMENTS: 6 MONTHS

**Investigator Global Assessment of i & ii) Pigment change iii) Atrophic scars iv) Hypertrophic/ Keloid scars**

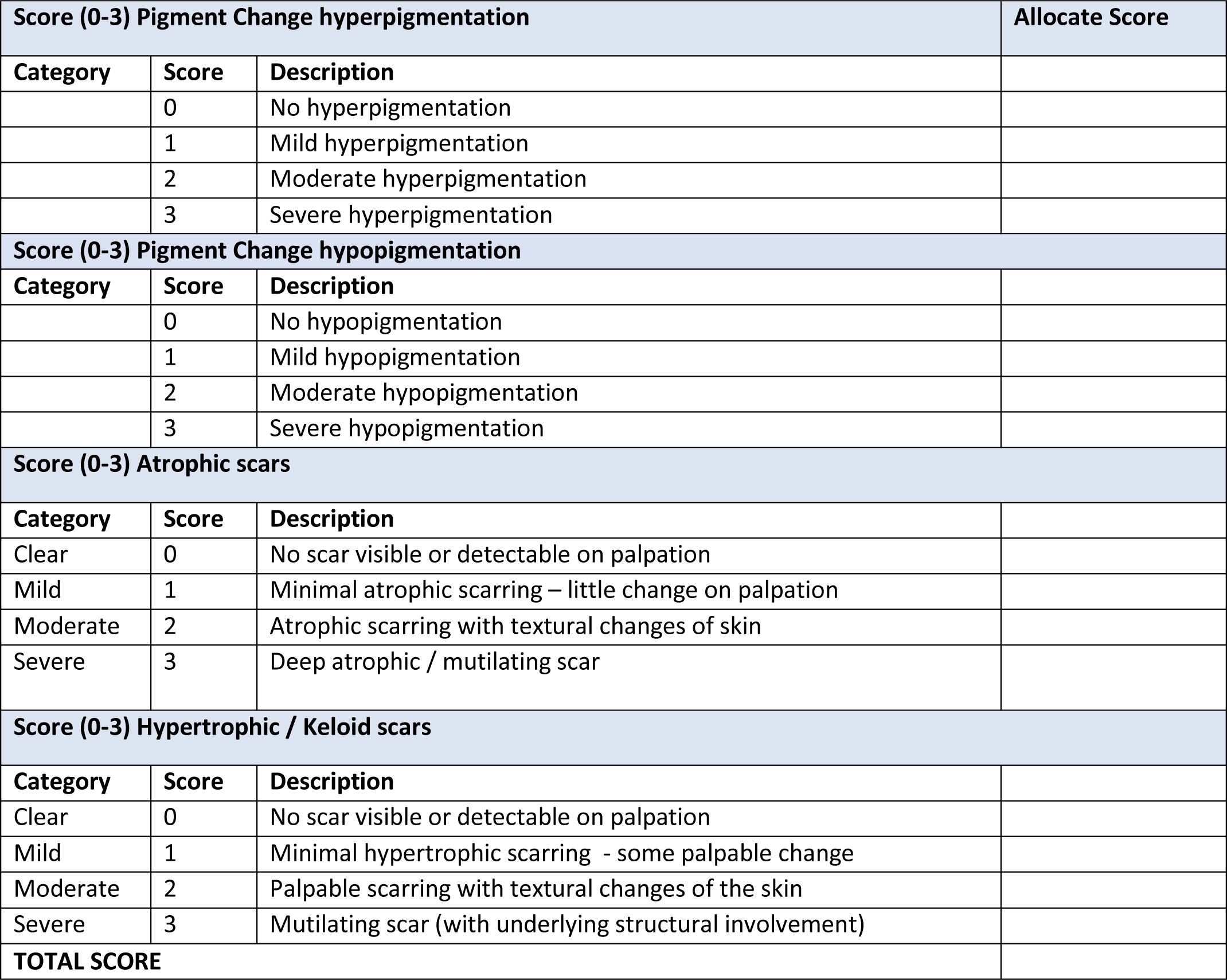

#### 9.6. HRQoL: 6 MONTHS

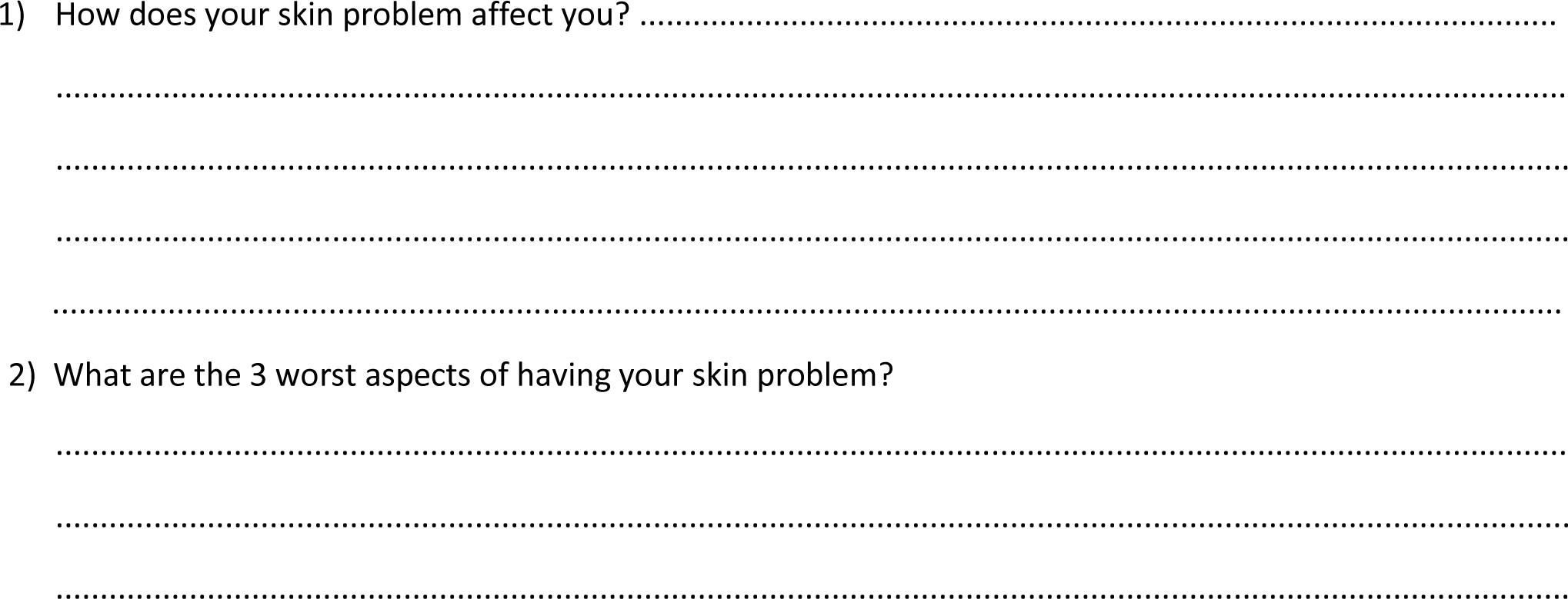

#### 9.7. SUMMARY OF SCORES: 6 MONTHS

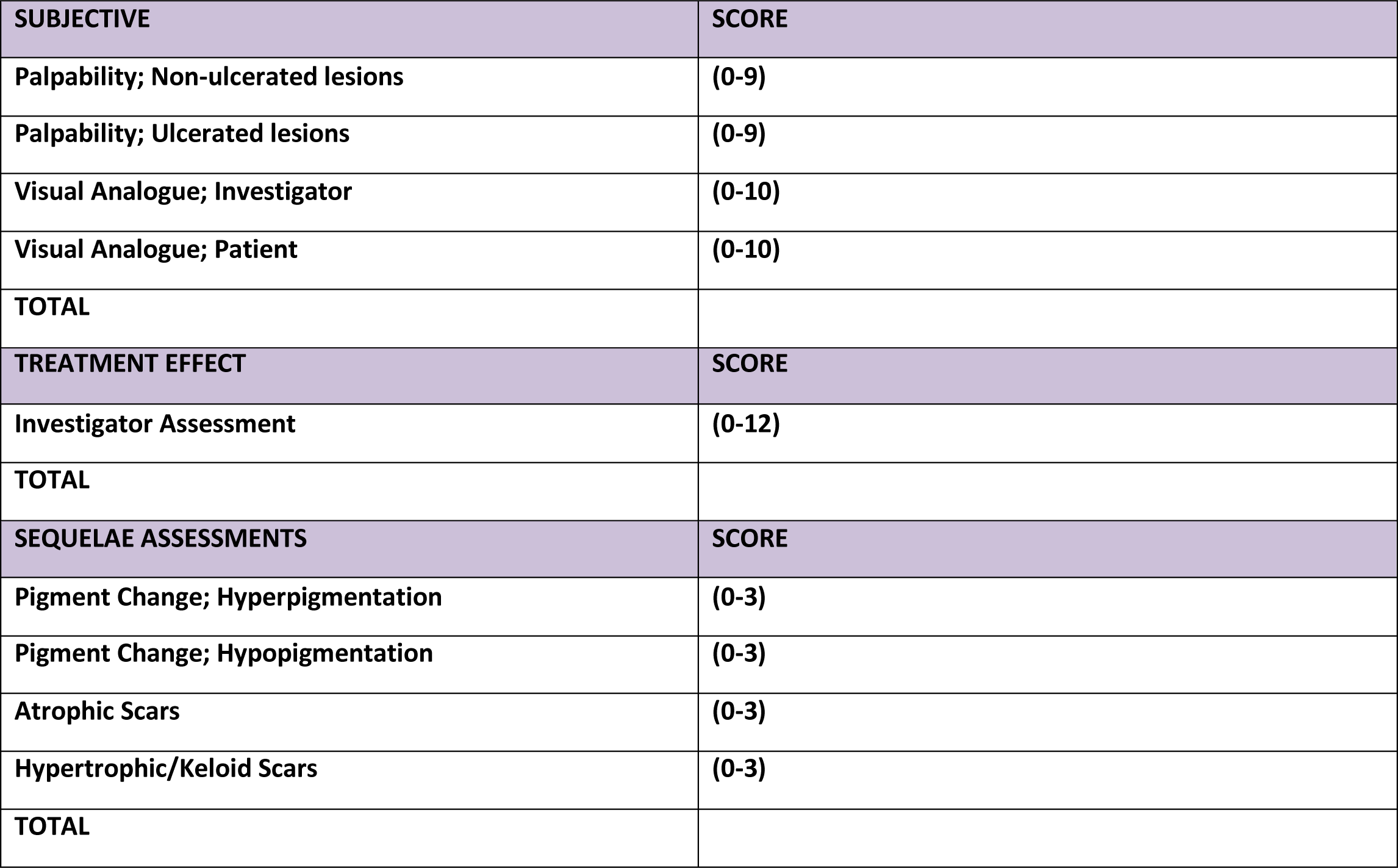

### 10. SUMMARY OF SCORES OVER TIME

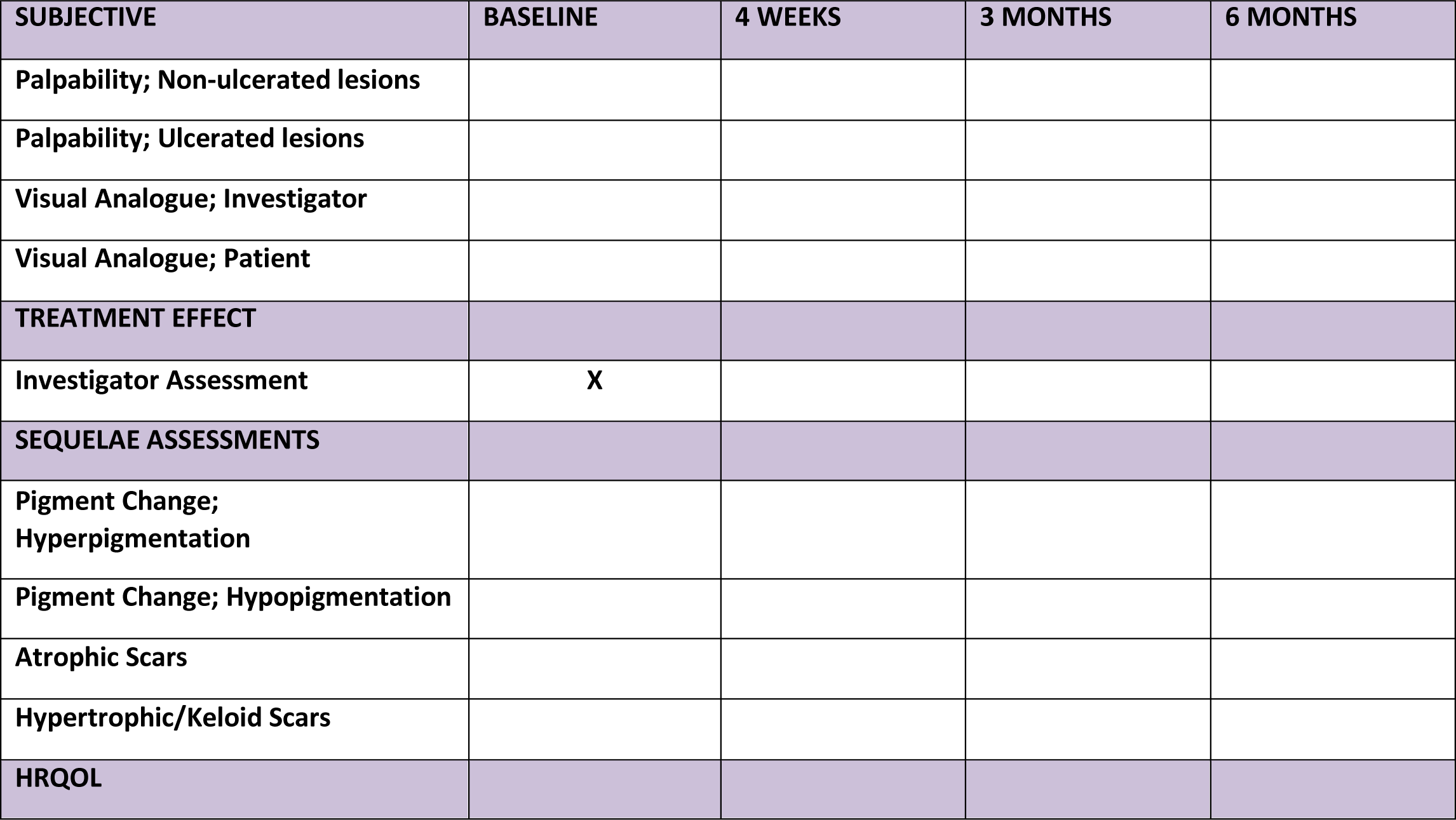

### 11. Drug therapy related to Leishmaniasis

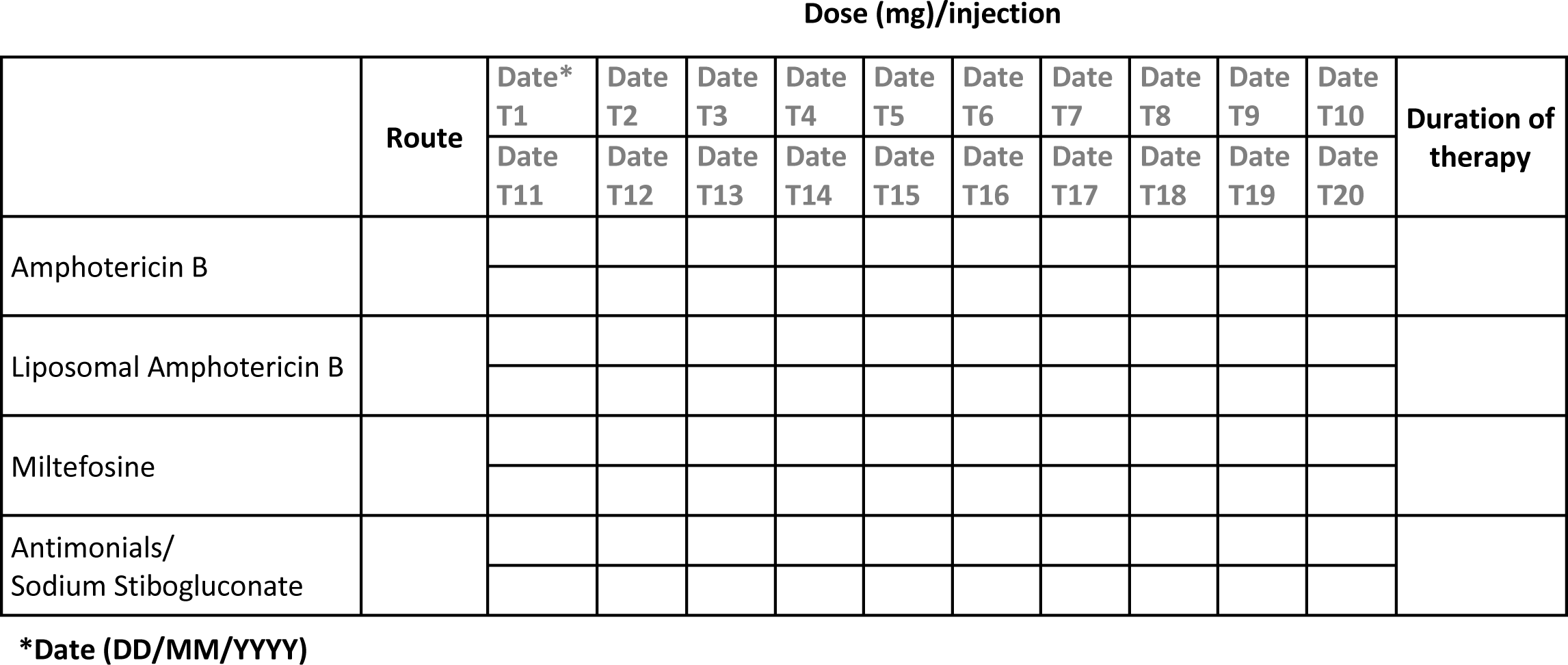

### 12. Cytokine profiles in dermal Lesions

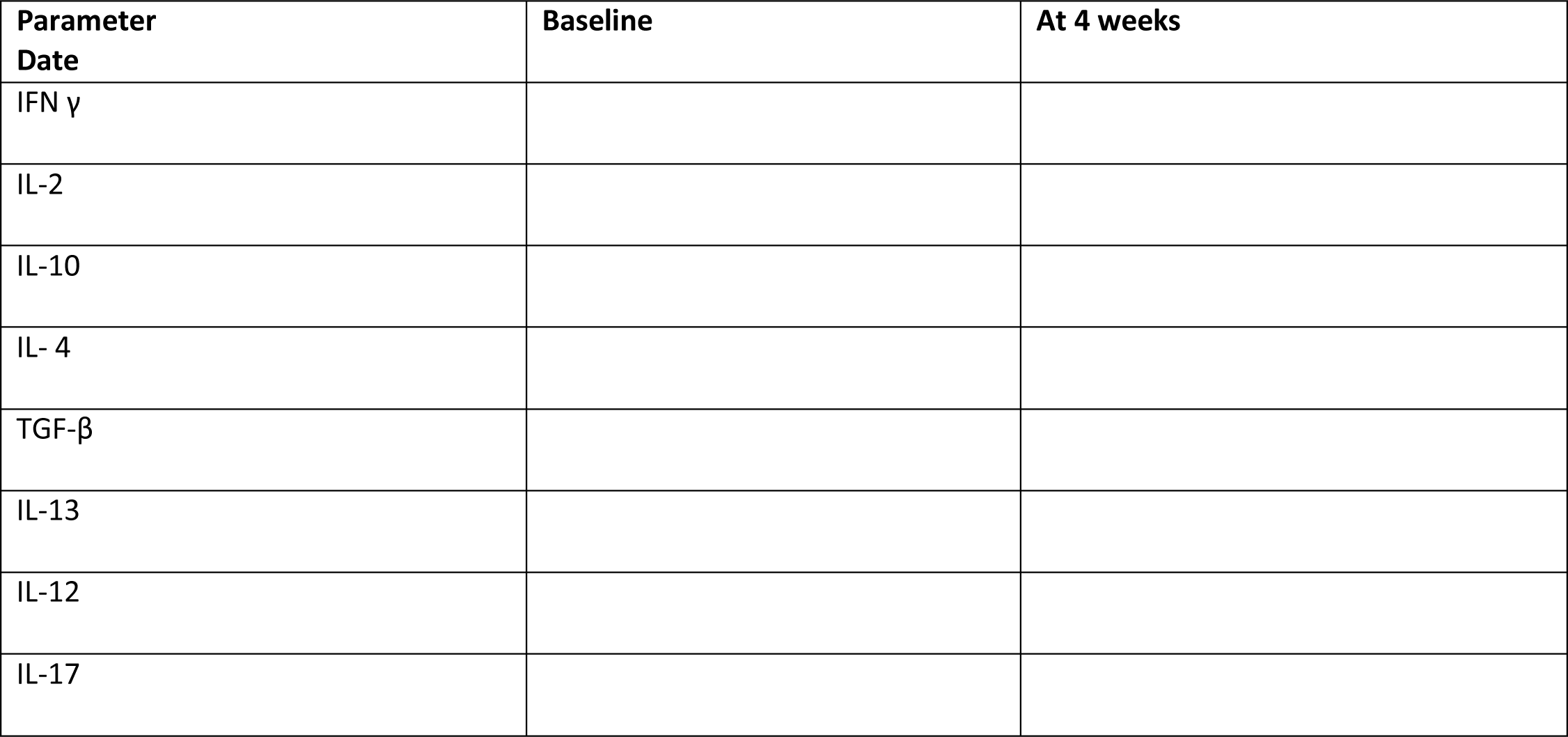

### 13. miRNA analysis of blood

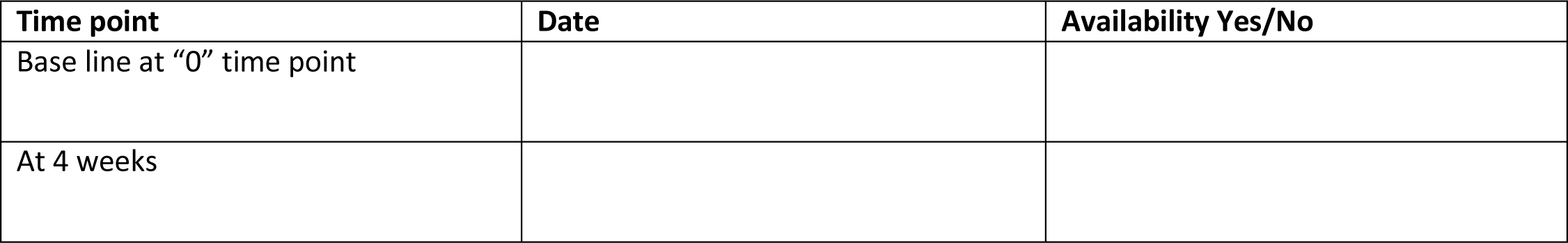

### 14. Complete Blood Count, Biochemical Parameters, Histopathology: include dates

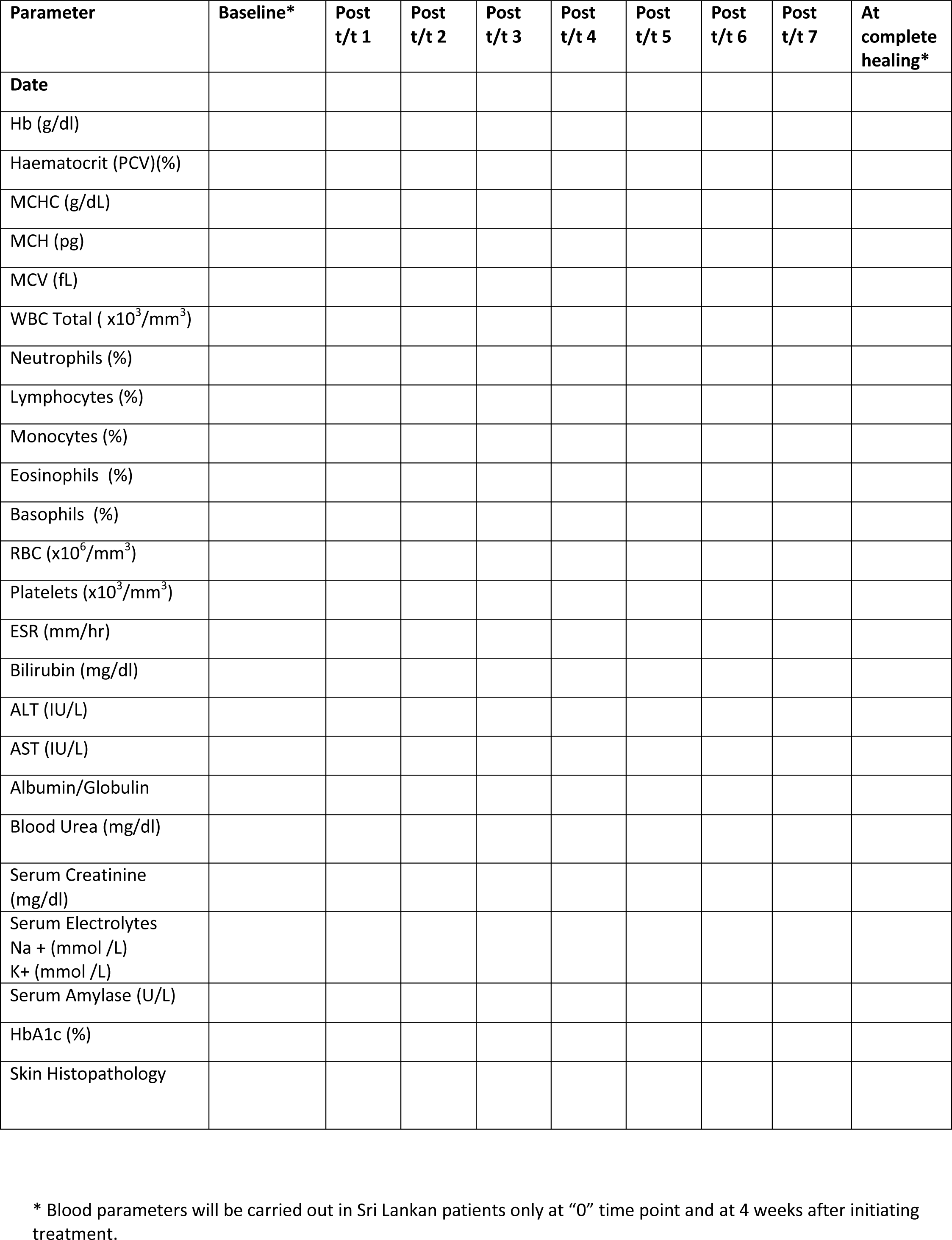

### 15. Final Impression (after completion of treatment) and additional comments

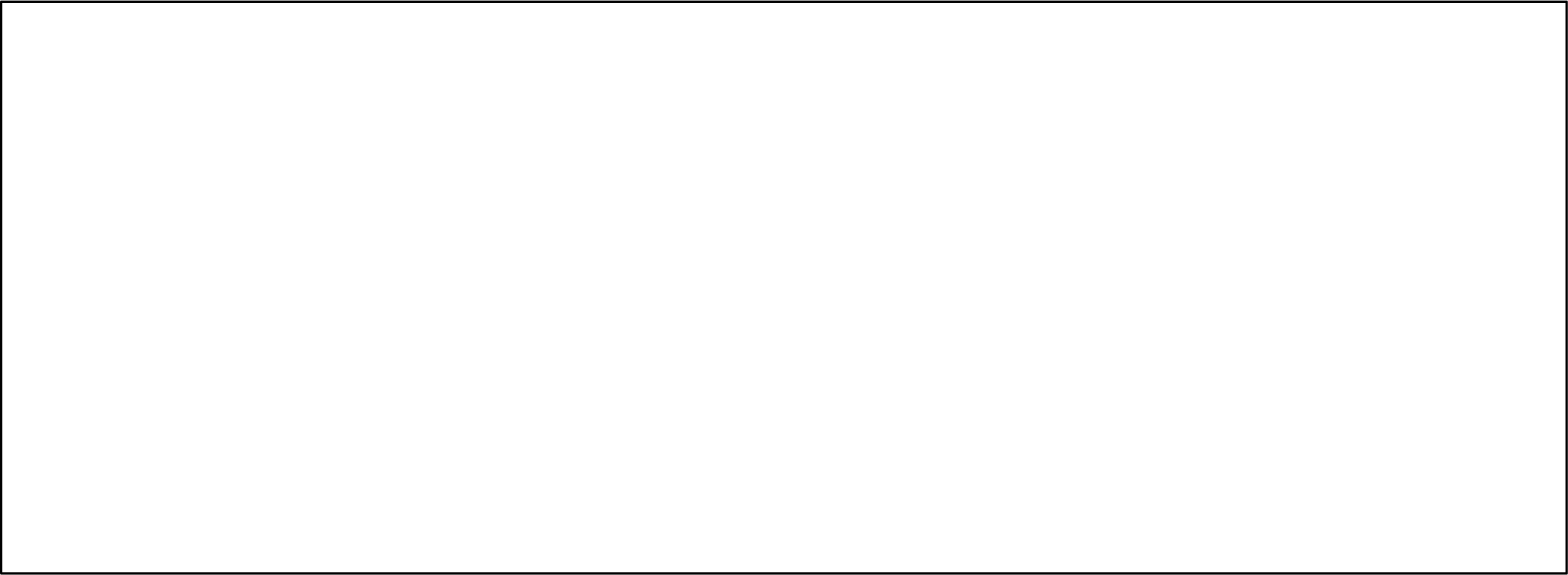

### 16. Investigator’s details: Name

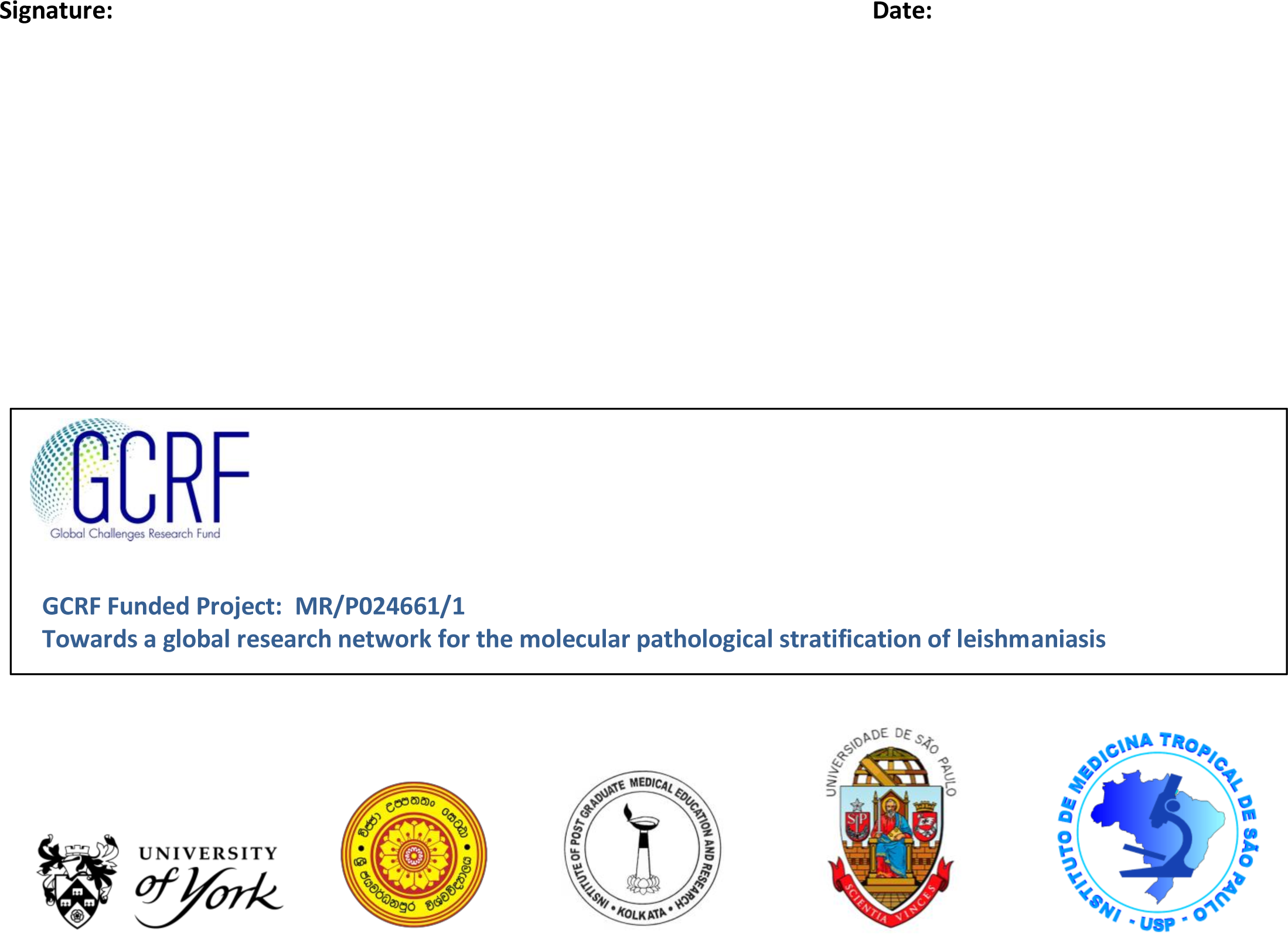

